# Functional Data Analysis: Transition from Daily Observation of COVID-19 Prevalence in France to Functional Curves

**DOI:** 10.1101/2021.09.25.21264106

**Authors:** Kayode Oshinubi, Firas Ibrahim, Mustapha Rachdi, Jacques Demongeot

## Abstract

In this paper we use the technique of functional data analysis to model daily hospitalized, deceased, ICU cases and return home patient numbers along the COVID-19 outbreak, considered as functional data across different departments in France while our response variables are numbers of vaccinations, deaths, infected, recovered and tests in France. These sets of data were considered before and after vaccination started in France. We used some smoothing techniques to smooth our data set, then analysis based on functional principal components method was performed, clustering using k-means techniques was done to understand the dynamics of the pandemic in different French departments according to their geographical location on France map and we also performed canonical correlations analysis between variables. Finally, we made some predictions to assess the accuracy of the method using functional linear regression models.

## 1. Introduction

### 1.1 Background and literature review

The modeling of COVID-19 pandemic across the globe has been approached using different techniques in mathematics and statistics by different researchers but the use of functional data analysis (FDA) has been done by few scientists. Functional data analysis is useful in many fields such as medical sciences, biology, statistical analysis and econometrics while several books like [1] have treated the theoretical aspects and methodology and more recently, researchers have dealt with FDA application to COVID-19 modeling [2-4].

The COVID-19 pandemic is still evolving in France as there has been three waves with possibility of a fourth wave due to a more contagious variant (Delta variant) which may lead to another lockdown following three lockdowns alongside with several non-pharmaceutical measures to mitigate the spread of the diseases. France has a total of 5,911,601 cases as at 22/07/2021, 111,554 deaths representing 2% of the total cases, 5,162,757 having recovered from the disease representing 98% of the total cases and 637,290 currently infected patients with 637,431 (99.9%) in mild condition while 859 (0.1%) are in critical condition.

Some researchers have worked on French public data on COVID-19 evolution in France which we shall point out few as there are many more. The robust phenomenological approach to France COVID-19 data was investigated by [14] and a new method to calculate the cumulative cases in France was proposed which illustrates the epidemic and endemic nature of the virus infection in France. [13] used methods like principal component analysis, generalized additive model and hierarchical ascendant classification to study the impacts of population age structure, epidemic spread and transmission mitigation policies on COVID-19 morbidity or mortality heterogeneity in France. [15] used ARIMA models with different parameters to forecast the spread of COVID-19 across nine countries in Europe, Asia and American continents and the study deduced that the method is useful for the prediction of the pandemic at different stages.

Some recent works use functional data analysis for the modelling of COVID-19 pandemic as follows: [2] applied functional data analysis to United States data by using FCPA (Functional Principal Component Analysis) and FCCA (Functional Canonical Correlation Analysis) tools and they finally use functional time series to fit the cumulative confirmed cases in the United States and make forecasts based on the dynamics of FPCA. [3] worked on the imputation of missing data of COVID-19 hospitalized and intensive care curves in Spain regions. They used function-on-function regression technique to estimate missing values and Canonical Correlation Analysis was performed to interpret the relationship between hospital occupancy rate and illness response variables. The shapes of an epidemic curve using functional data analysis to characterize COVID-19 in Italian regions and their association with mobility, positivity, socio-demographic structure and environmental covariates was worked on by [4]. The researchers have used different methods of functional data analysis like function-on-function regression techniques, clustering methods and smoothing techniques for the functional data considered.

### 1.2 Time series and curve fitting

Figure 1a gives the time series of recent daily cases of COVID-19 in France which shows stationarity with rolling values (window=12) appearing to be varying slightly. Also, the statistics are smaller than the 5% (p-value = 0.02) critical values so we can say with 95% confidence that this is a stationary series. Also, in Figure 1c we plotted three French departments (Nord, Paris and Essonne) with more prevalent COVID-19 hospitalization cases and Figure 1d shows the fitness curve of two of the French departments (Paris and Seine-Maritime) while all departments have root mean square error in the interval 0.51 ≤ *RMSE* ≤ 17.38 with Essonne department having the highest RMSE and Lozère department having the lowest RMSE. We present other RMSE values in Table 1. We present also (Figure 1b) a deep learning forecasting result using Gated Recurrent Units (GRU) for France data between the beginning of the pandemic in France till September 3 2021 by training 80% of the data and testing 20%. The predicted cases curve values decline over the whole-time. In Appendix B, we present the spectral analysis of the time series in Figure 15 in order to study the periodicity of the new cases of COVID-19 in France and to present a smoothed version without noise of the data.

**Figure 1.**
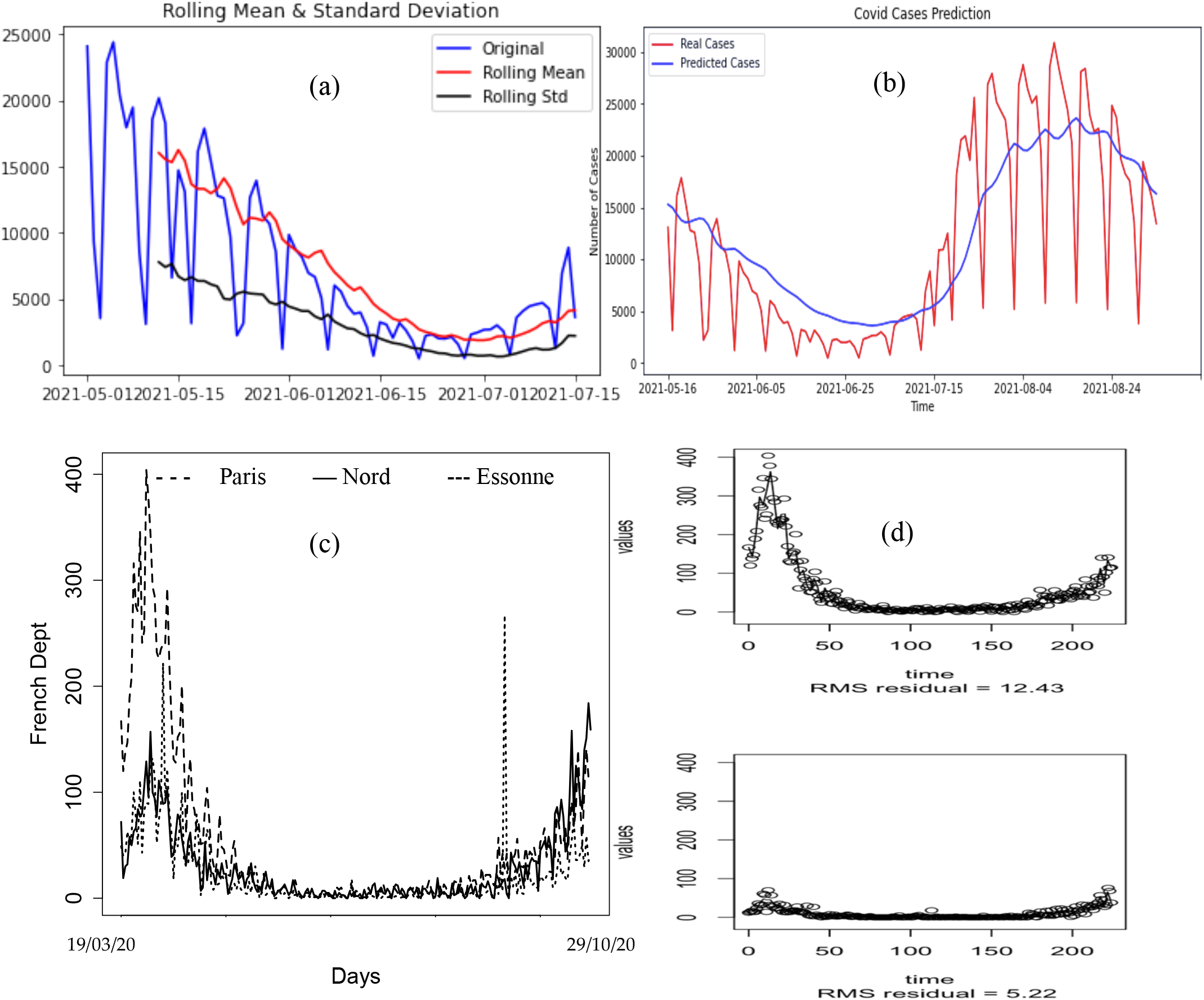
(a) Time series modelling of daily new cases between 01/05/2021 to 15/07/2021 in France. (b) GRU deep learning forecasting method for daily new cases between 25/02/2020 to 03/09/2021 in France. (c) Daily hospitalization cases in three French departments: Nord, Paris and Essonne. (d) Fit curve for hospitalization cases in Paris and Seine-Maritime.

**Table 1.** RMSE confidence interval for all French departments for the fitness curve of the four functional data

|  | RMSE before vaccination started | RMSE after vaccination has started |
| --- | --- | --- |
| Hospitalized | $0.51 \leq RMSE \leq 17.38$ | $1.00 \leq RMSE \leq 18.00$ |
| ICU | $0.05 \leq RMSE \leq 2.60$ | $0.35 \leq RMSE \leq 5.20$ |
| Daily return home | $0.25 \leq RMSE \leq 12.40$ | $1.10 \leq RMSE \leq 17.50$ |
| Daily deceased | $0.04 \leq RMSE \leq 4.52$ | $0.32 \leq RMSE \leq 4.10$ |

The aim of this paper is to model the prevalence of the virus in France by using several functional techniques like FCCA, K-means clustering and FPCA and to finally make some predictions about the evolution of the disease in France. The analysis was done using both Python and R packages. We considered as functional variables numbers of ICU cases, daily deceased, daily return home and hospitalization which are given as *X*_1_, *X*_2_, *X*_3_ and *X*_4_. Our response variables given as *Y*_1_, *Y*_2_, *Y*_3_, *Y*_4_, *Y*_5_ and *Y*_6_ are numbers of recovery, deaths, infected, vaccination, vaccination per 1000 population and number of tests. We collated data from [10], [11] and [12]. The paper is divided as follows: in Section 2 we describe the various smoothing methods employed in the analysis of the shapes of the functional data used and in Section 3 we present the functional principal components analysis results and their interpretation to the dynamics of COVID-19 prevalence in French departments, Section 4 is dedicated to the result of canonical correlation of the variables. In Section 5 we present the clustering result using K-means method and how it appears on the map of France, In Section 6 we made some predictions for some response variables and also performed the function-on-function linear regression and finally in Section 7 we opened up some perspectives and gave conclusion of the analyses.

## 2. Data smoothing

The first step in analyzing functional data is to smooth the curves. In this Section we use different smoothing techniques which we shall illustrate and give some basic explanation of the techniques we deployed for smoothing our functional data. We plotted the mean of the data set and the cross-sectional mean, which corresponds to the karcher-mean under the 𝕃^2^ distance [8]. We used the *elastic_mean(fd)* and *fd.mean* tool in Python to do the plotting of Figure 2.

**Figure 2.**
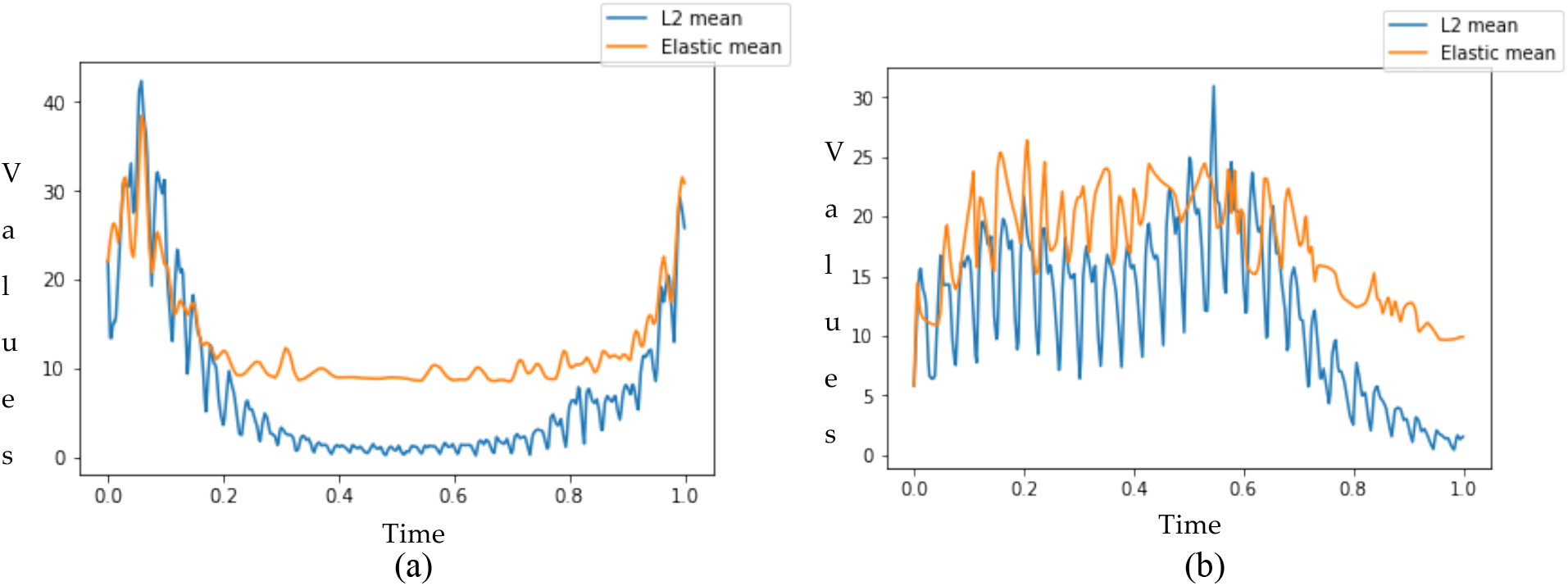
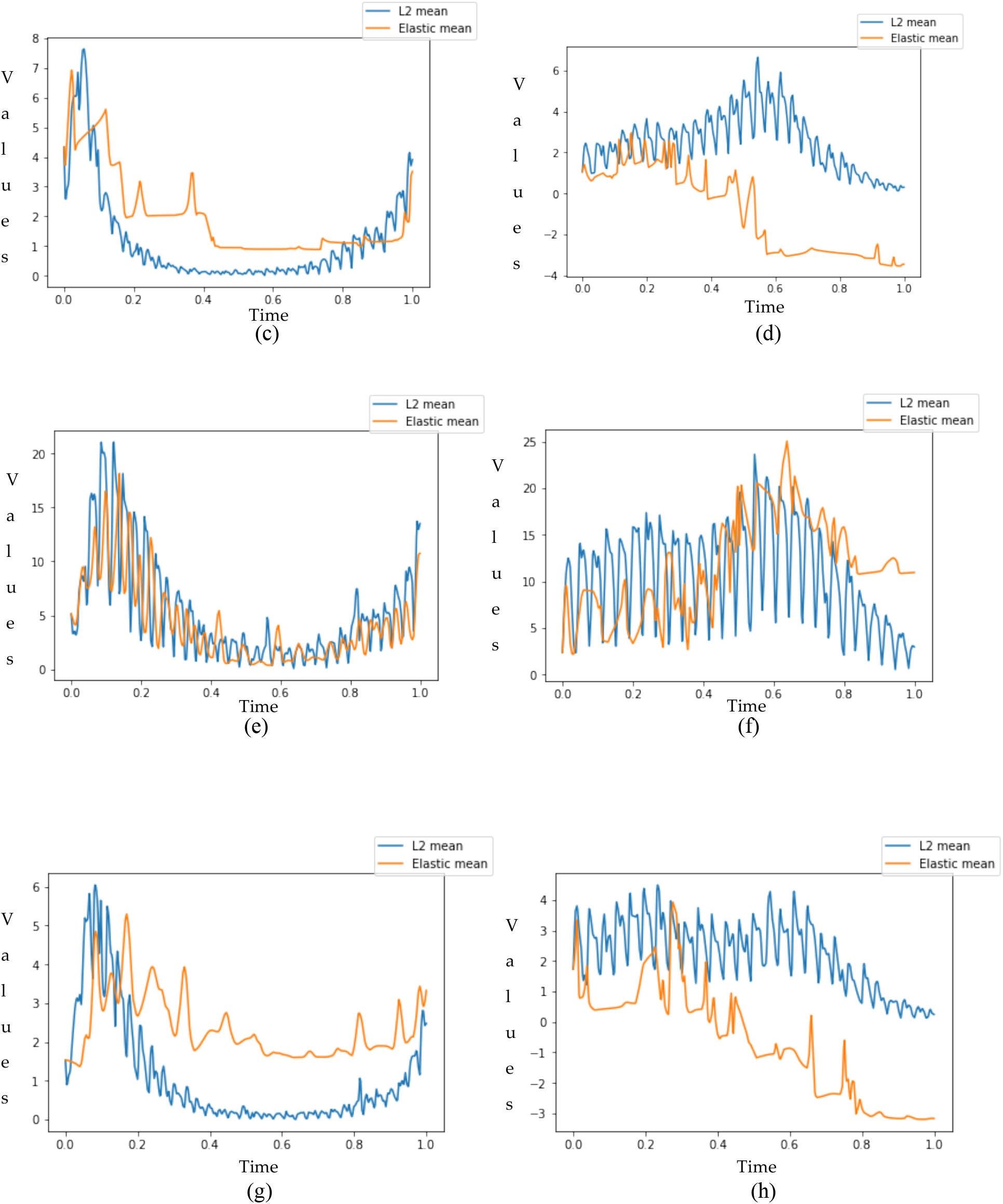
L2 (blue) and elastic (yellow) means of the functional data: (a) hospitalized cases, (b) hospitalized when vaccination has started, (c) ICU cases, (d) ICU cases when vaccination has started, (e) daily return home, (f) daily return home when vaccination has started, (g) daily deceased and (h) daily deceased when vaccination has started.

We observed that the elastic mean better captures the geometry of the curves compared to the standard L2 mean for some of the functional data set we considered, since it is not affected by the deformations of the curves. This phenomenon can be seen in Figures 2a, 2b, 2c, 2e, 2f and 2g, Figures 2d and 2h showing a bad shape for elastic mean.

### B-Spline smoothing technique

B-spline technique is one of the tools used in smoothing a functional data and this can be done by changing the number of elements (n = 2,3,4,…) in the basis functions [9]. Sometimes one can use the Fourier basis for the functions to further see the variations in the curves. We give a mathematical expression on the basis functions below:

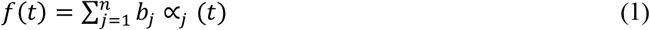

For this analysis we choose n= 7 as our number of elements and the tool in Python named *basis.BSpline* was used to perform the plotting of the functional data. The result of this smoothing technique can be seen in Figure 3.

**Figure 3.**
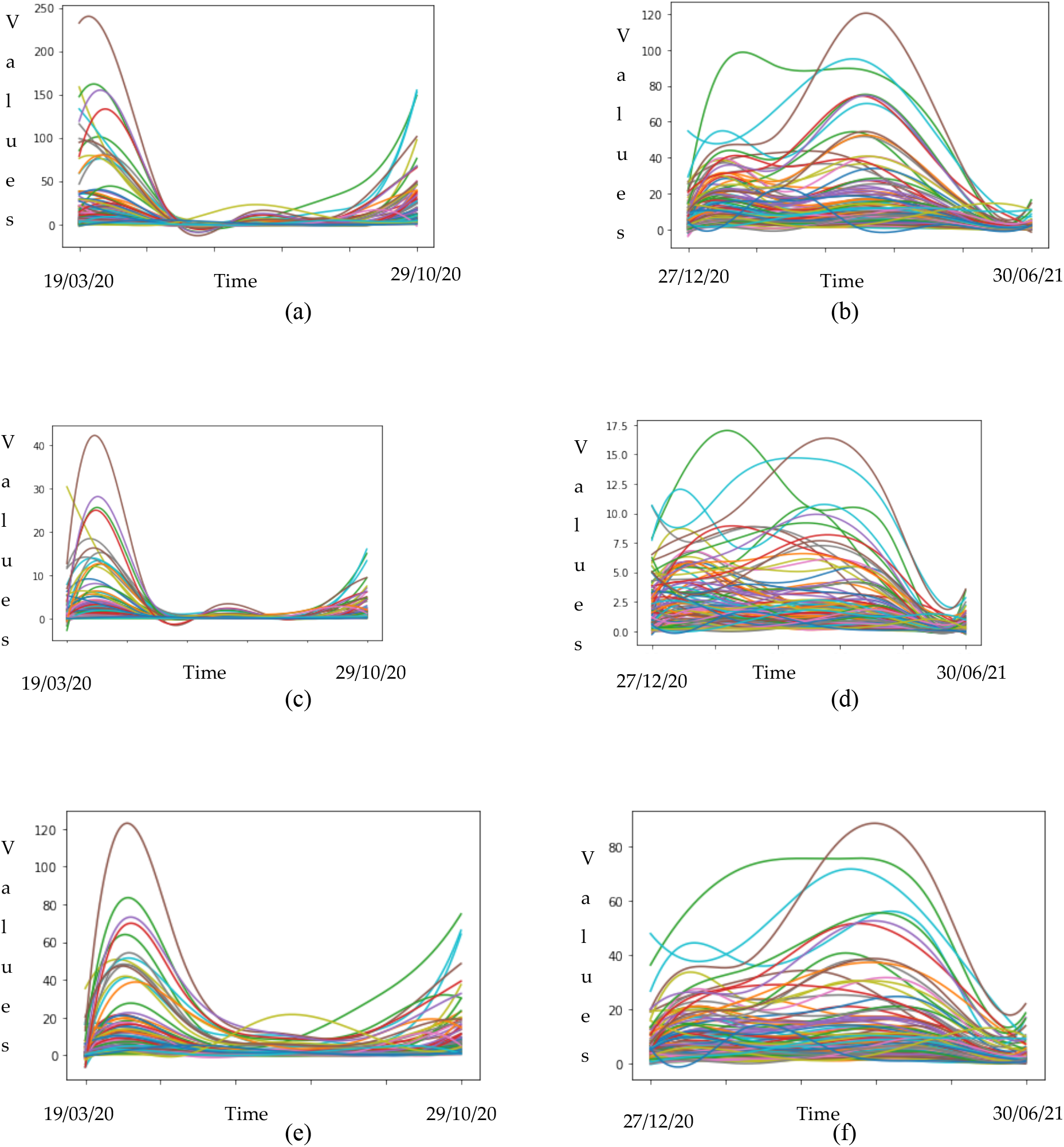
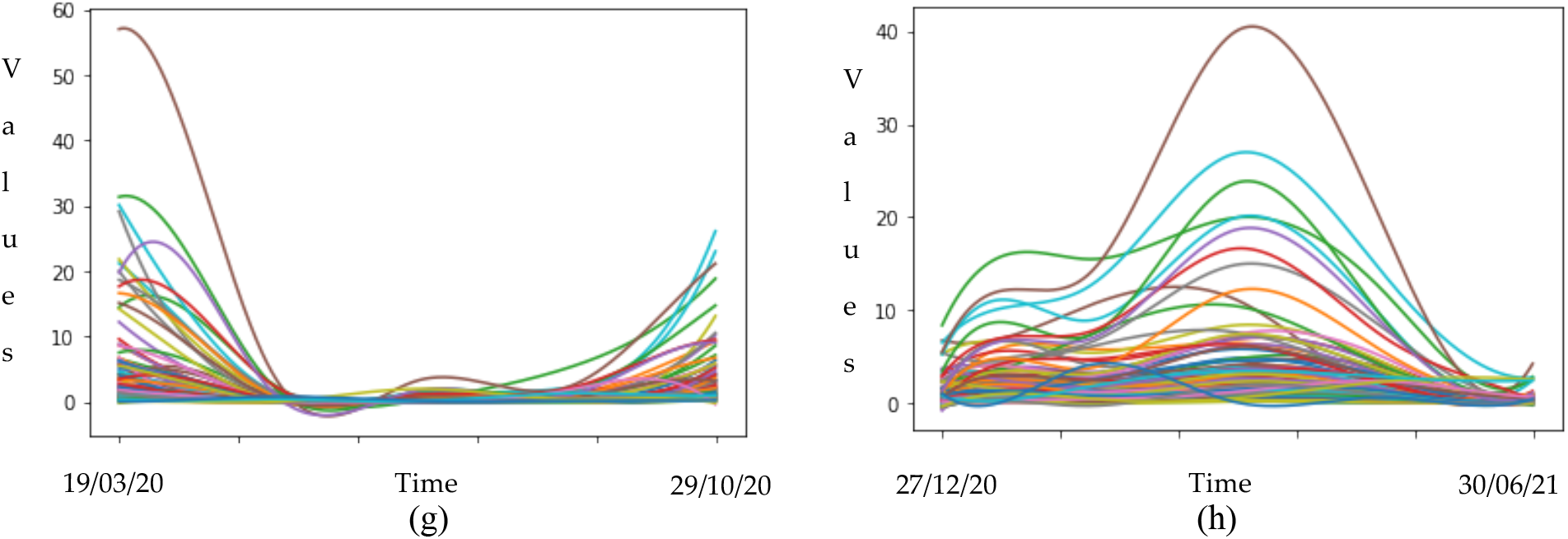
Smoothed curves for the shape of COVID-19 epidemic in all departments in France: ((a) hospitalized cases, (b) hospitalized when vaccination has started, (c) daily deceased, (d) daily deceased when vaccination has started, (e) daily return home, (f) daily return home when vaccination has started, (g) ICU cases and (h) ICU cases when vaccination has started).

In Appendix A Figure 14, we presented the correlation coefficient between all the departments in France based on the functional data in consideration, in order to see how well our data is well correlated between the departments and it was observed that there is a high correlation between various departments with except in few cases where we observed low correlation as we can see in the contour plots presented in Figures 14a to 14h.

### Smooth interpolation and monotone cubic spline interpolation

We used spline interpolation of order 3 and then smooth the interpolation using the smoothness parameter equal to 1.5 in the cubic spline smoothing. This technique is demonstrated using *interpolation* and *smoothness_parameter* package in Python. We also use the monotone technique and a piecewise cubic Hermite interpolating polynomial (PCHIP) using a Python package called *monotone*. We present some of the results on Figure 4.

**Figure 4.**
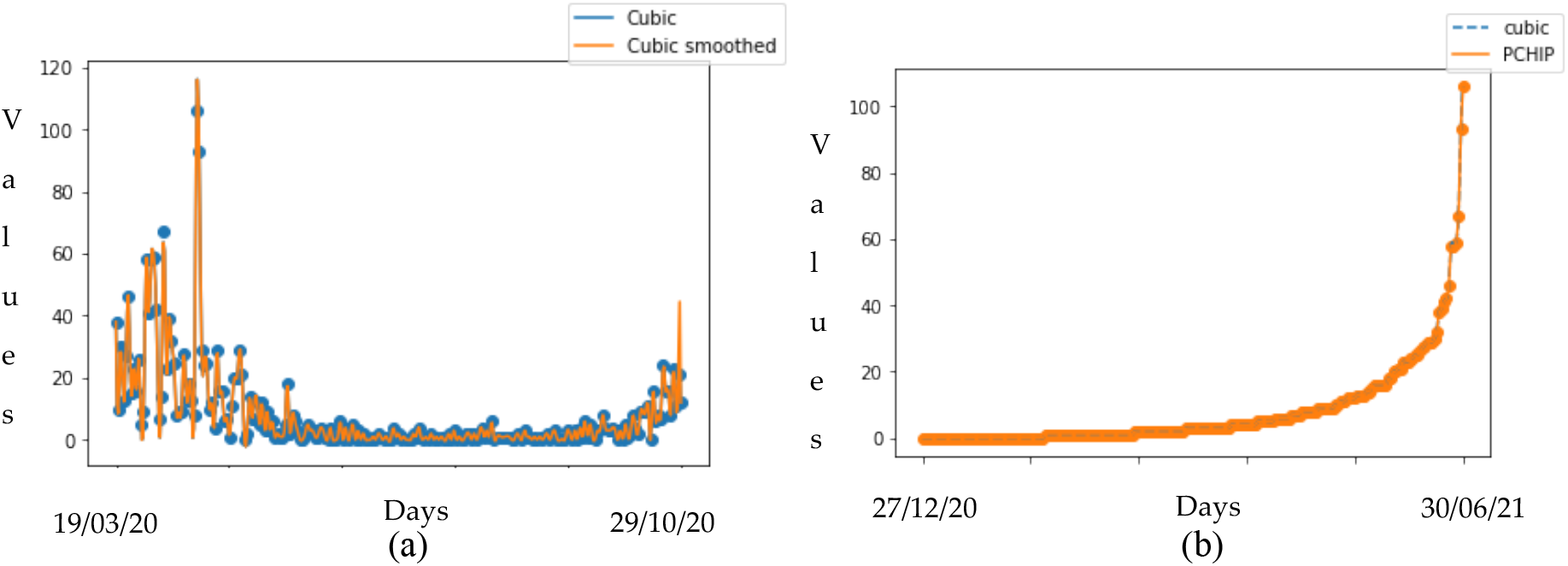
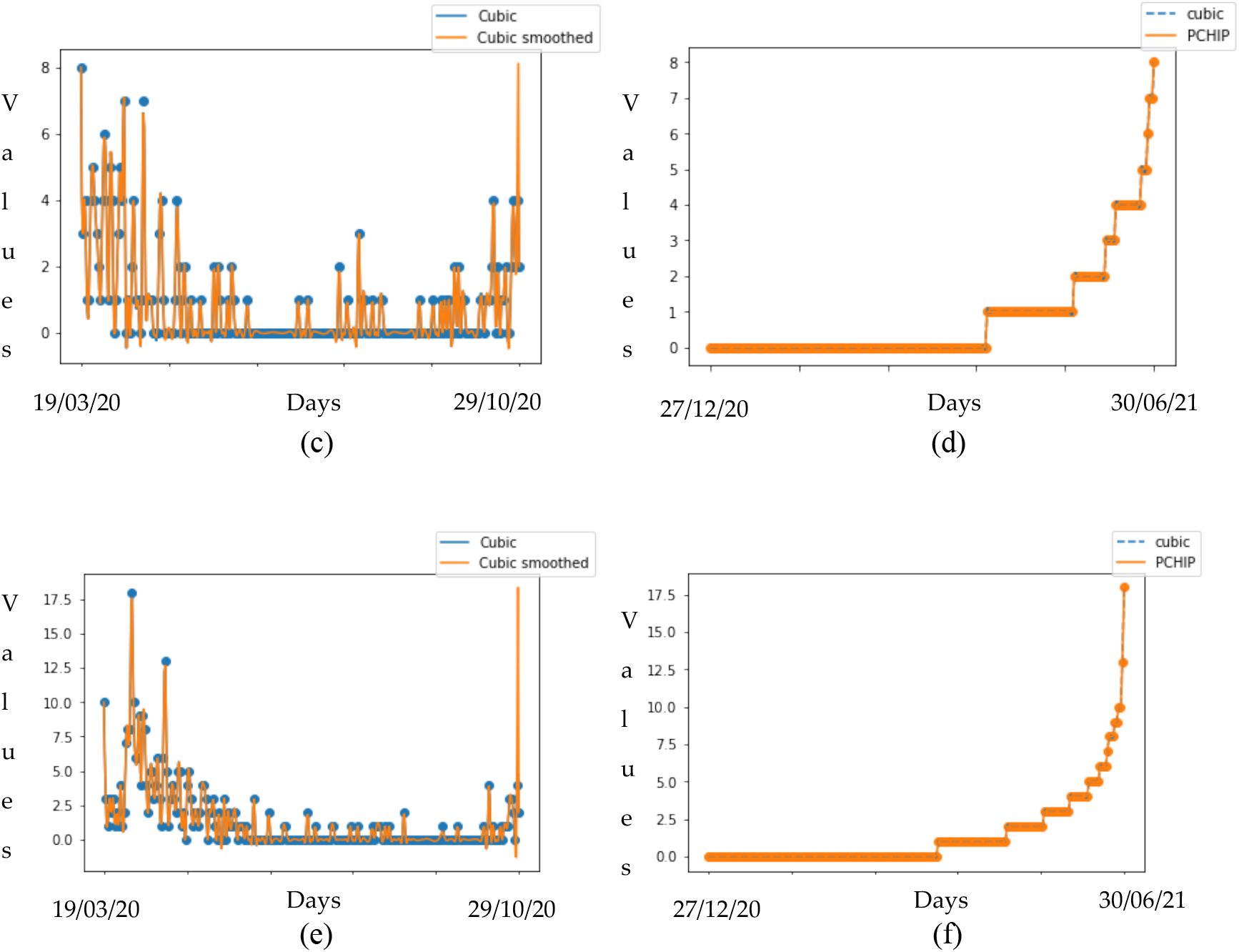
(a) Hospitalized cases interpolation smoothing, (b) hospitalized monotone smoothing, (c) ICU cases interpolation smoothing, (d) ICU cases monotone smoothing, (e) daily deceased interpolation smoothing and (f) daily deceased monotone smoothing.

From graphs presented in Figure 4 one can deduce that cubic spline smoothing curves exhibit oscillations and oscillations are important to know the low and high data thresholds in case the consecutive data points experience a significant change in slope. We also observed that PCHIP is smooth and non-oscillatory despite some sharp increase as the U-shape of the curve deepens.

### Kernel smoothing

We also performed Kernel smoothing to show how cross validation score varies over a range of different parameters used in the smoothing methods. The essence of this section is to estimate the smoothing parameter *h* that better represents the functional data and this smoothing parameter was selected by generalised cross-validation criteria. The non parametric method of smoothing for the functional data is based on smoothing matrix M given below:

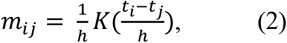

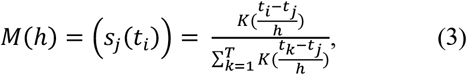

where K() is the Kernel function.

We plotted on Figure 5 the smoothed curves of the functional data set for three different smoothing methods and also show the scores through generalised cross-validation (GCV) for these different smoothing methods. The results show a comparable behavior of these scores by varying the smoothing parameter *h*.

**Figure 5.**
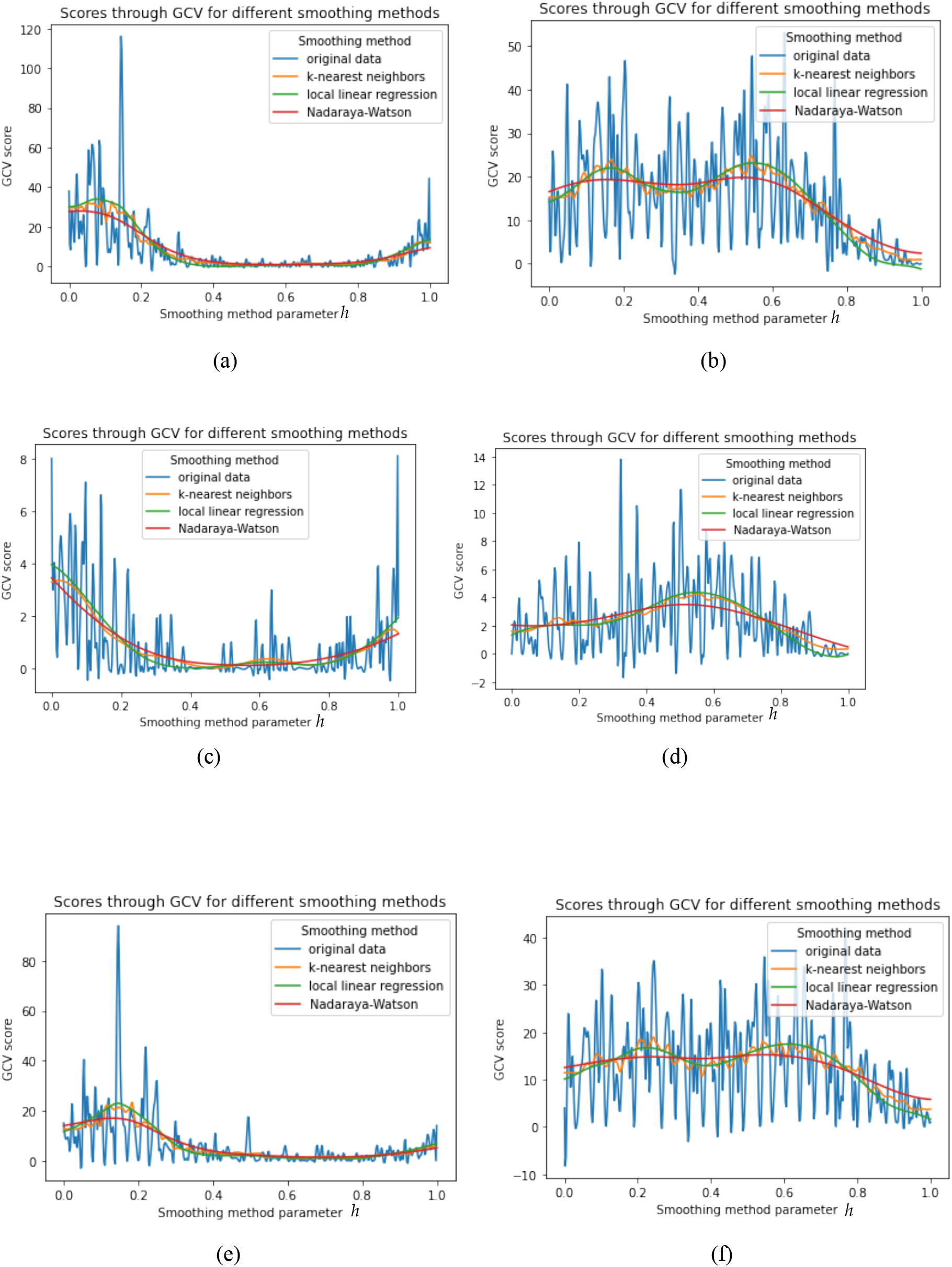
Kernel smoothing method for (a) hospitalized cases, (b) hospitalized when vaccination has started, (c) ICU cases, (d) ICU cases when vaccination has started, (e) daily return home and (f) daily return home when vaccination has started.

## 3. Functional principal component analysis (FPCA)

Principal component analysis is a dimension reduction analysis tool in multivariate statistics while functional principal component analysis (FPCA) is a dimension reduction with high variance in functional data analysis [6], [7]. Let *x*_*i*_(*t*) be a given set of functions and let *α* be a weight, FPCA is computed as follows:

- It finds the principal component weight function *α*_1_(*t*) for which the principal component score is given by

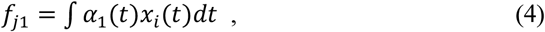
- while maximizing 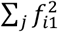 is subjected to

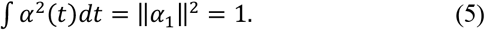
- Next, the weight function *α*_2_(*t*) is computed and the principal component score maximizes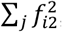, and is subject to the constraint ‖*α*_2_‖^2^ = 1 and to the additional constraint

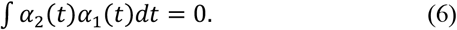
- Then, the process is repeated for as many iterations.

In our analysis we used a tool called *pca.fd* for the principal component analysis. We present in this Section the 4 PCs values plot throughout the days considered and the principal component scores plot for all the different departments providing functional data being before vaccination started and during vaccination.

### Functional PCs

- Hospitalization cases In Figure 6a we observed that PC1 peaked in the early days of the pandemic between February and March 2020 and then there was a decline after about 50 days becoming stationary till day 150 possibly due to mitigation measures promulgated during this period. The same phenomenon has been observed for PC 2. In Figure 6a, PC 4 shows a sinusoidal shape, peaked at day 100 which is around June 2020 with least values at day 30 and day 180 which are respectively in March and August 2020. Figure 6b shows the same sinusoidal shape for PC 4 and same shape for PC 3 but with a drift in the observation with a difference between the dynamics of hospitalization cases before and after vaccination has started in France. PC 1 in Figure 6b shows a decline across the infective period which may be due to the aggressive vaccination campaign in the country.
- ICU cases In Figure 6c we observed that from day 50 (around April 2020) till day 150 (around July 2020), the PC 1 value which is the major PC is stable throughout this period of various confinement measures in France and all PCs tend to show increasing behavior after the confinement measures have been relaxed and in Figure 6d, PC 1 has strictly positive values while PC 2, PC 3, PC 4 show negative values between February to June 2021.
- Daily return home In Figure 6e PC 1 peaked with a positive value at the beginning of the pandemic in France which validates the percentage of recovery as presented in the introduction Section while PC1 in Figure 6f shows a positive decline across the days considered, with a disparity between the period of vaccination and without vaccination.
- Daily deceased On the y-axis of Figures 6g and 6h, we observe that this is the only result with low values for the PCs because the deaths due to COVID-19 in France remain at a low level, while all PCs show almost the same pattern as that observed in previously for the other variables.

**Figure 6.**
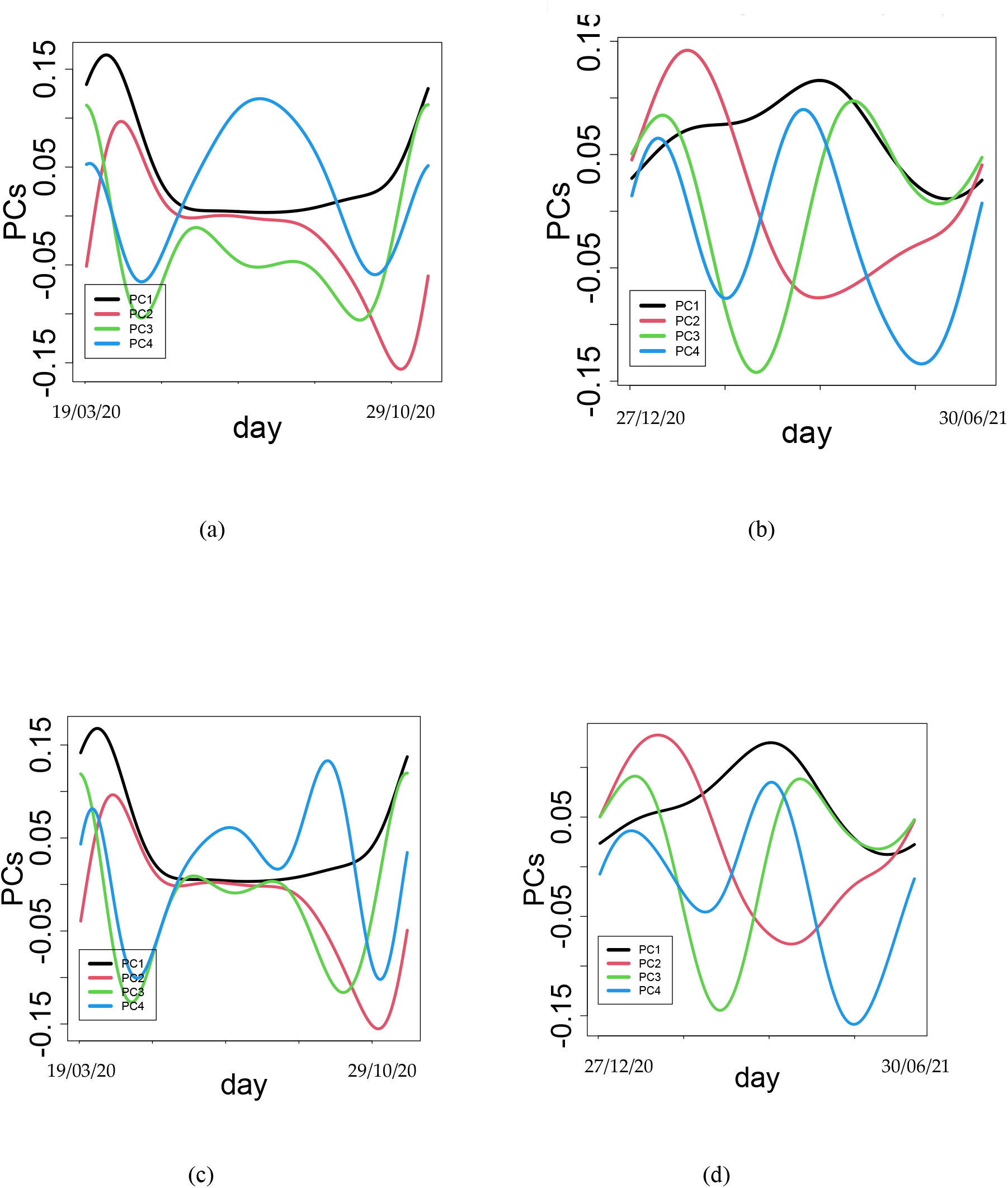
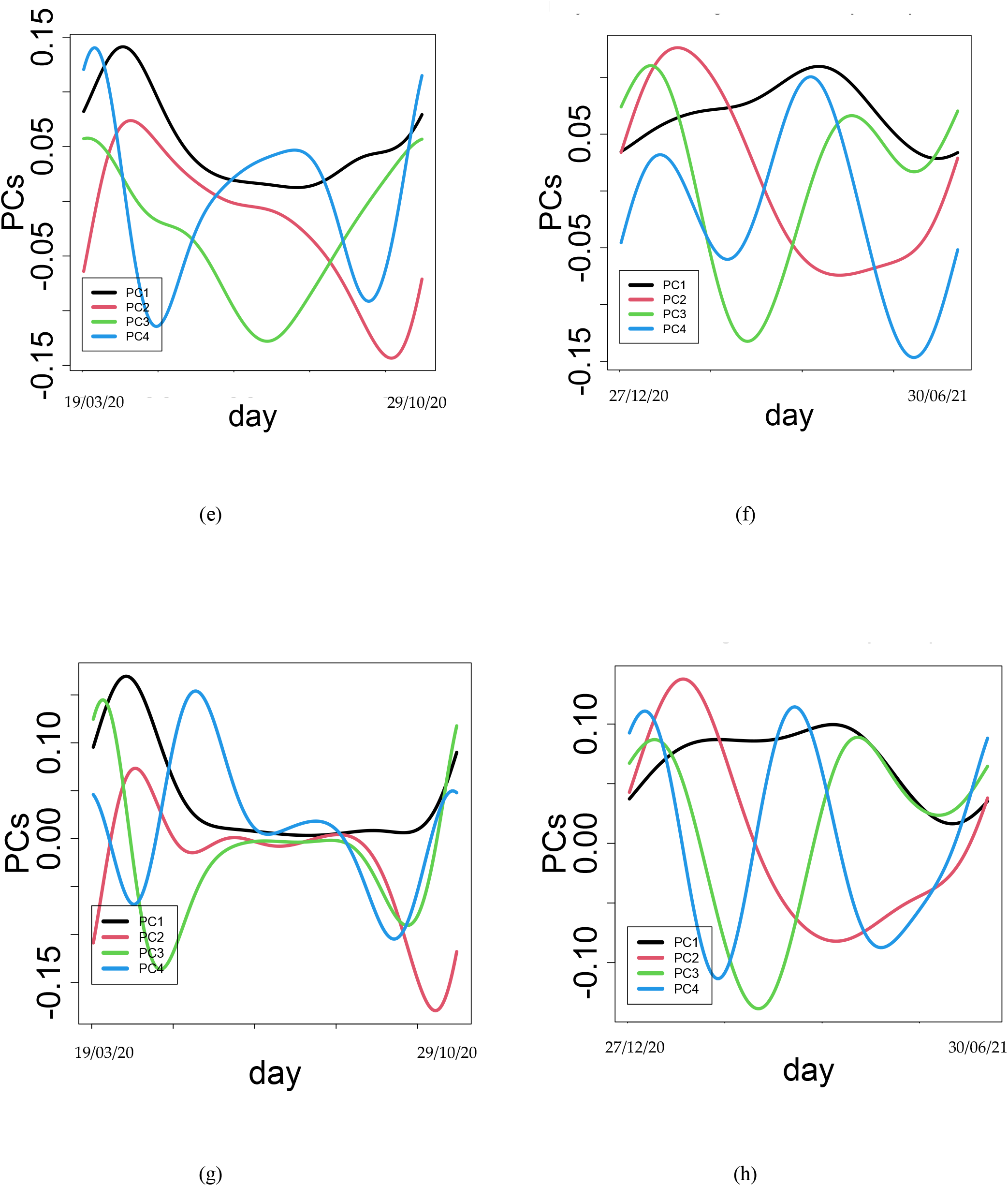
Functional PCs for different functional data before the start of vaccination (19/03/2020 to 29/10/2020) and when vaccination has started (27/12/2020 to 30/06/2021): (a) hospitalized cases, (b) hospitalized when vaccination has started, (c) ICU cases, (d) ICU cases when vaccination has started, (e) daily return home, (f) daily return home when vaccination has started, (g) daily deceased and (h) daily deceased when vaccination has started.

In Table 2, we present the PCs variance proportion and we observe that PC 1 is the most important PC.

**Table 2.** PCA variance proportion for 4 PCs

|  | Before vaccination started |  |  |  | After vaccination has started |  |  |  |
| --- | --- | --- | --- | --- | --- | --- | --- | --- |
|  | PC1 | PC2 | PC3 | PC4 | PC1 | PC2 | PC3 | PC4 |
| Hospitalized | 0.945 | 0.039 | 0.008 | 0.005 | 0.938 | 0.041 | 0.012 | 0.004 |
| ICU | 0.960 | 0.028 | 0.009 | 0.001 | 0.962 | 0.023 | 0.008 | 0.004 |
| Daily return home | 0.925 | 0.045 | 0.015 | 0.007 | 0.953 | 0.030 | 0.009 | 0.004 |
| Daily deceased | 0.965 | 0.017 | 0.013 | 0.003 | 0.914 | 0.055 | 0.016 | 0.010 |

### Functional principal component scores and clusters

We will now focus more on the departments in which the pandemic is more prevalent and also on PC 1 and PC 2, neglecting the other PCs. In Figure 7a, the Paris department (code number 75) and Nord department (code number 59) have a positive score in PC 1 and negative score in PC 2 while the Essonne department is positive in both PCs. In Figure 7b, the Paris department and Essonne department (code number 91) are negative in both PCs while the Nord department is positive in PC 2 with the highest score and negative in PC1. In Figure 7c, Nord and Essonne departments are negative in PC 2 but positive in PC 1 while the Paris department is positive in both PCs. The Paris department and Essonne department are negative in both PCs in Figure 7d while the Nord department is positive in PC 2 and negative in PC1. In Figure 7e, Paris and Nord departments have positive scores in both PCs while the Essonne department is negative in PC2 and positive in PC 1. Nord department has the highest positive score in PC 1 for Figure 7f and negative for PC1, Paris department is positive in PC 2 and negative in PC while Essonne department is negative in both PCs. The Paris department has the highest positive score in PC 1 and negative in PC 2 in Figure 7g, Nord department is positive in both PCs while Essonne department is negative in PC 2 but positive in PC 1. Finally, in Figure 7h while Nord department is positive and highest in PC 2, Paris department is the lowest with negative score in PC 2. Both departments are negative in PC 1. The Essonne department is positive in PC 2 but negative in PC 1.

**Figure 7.**
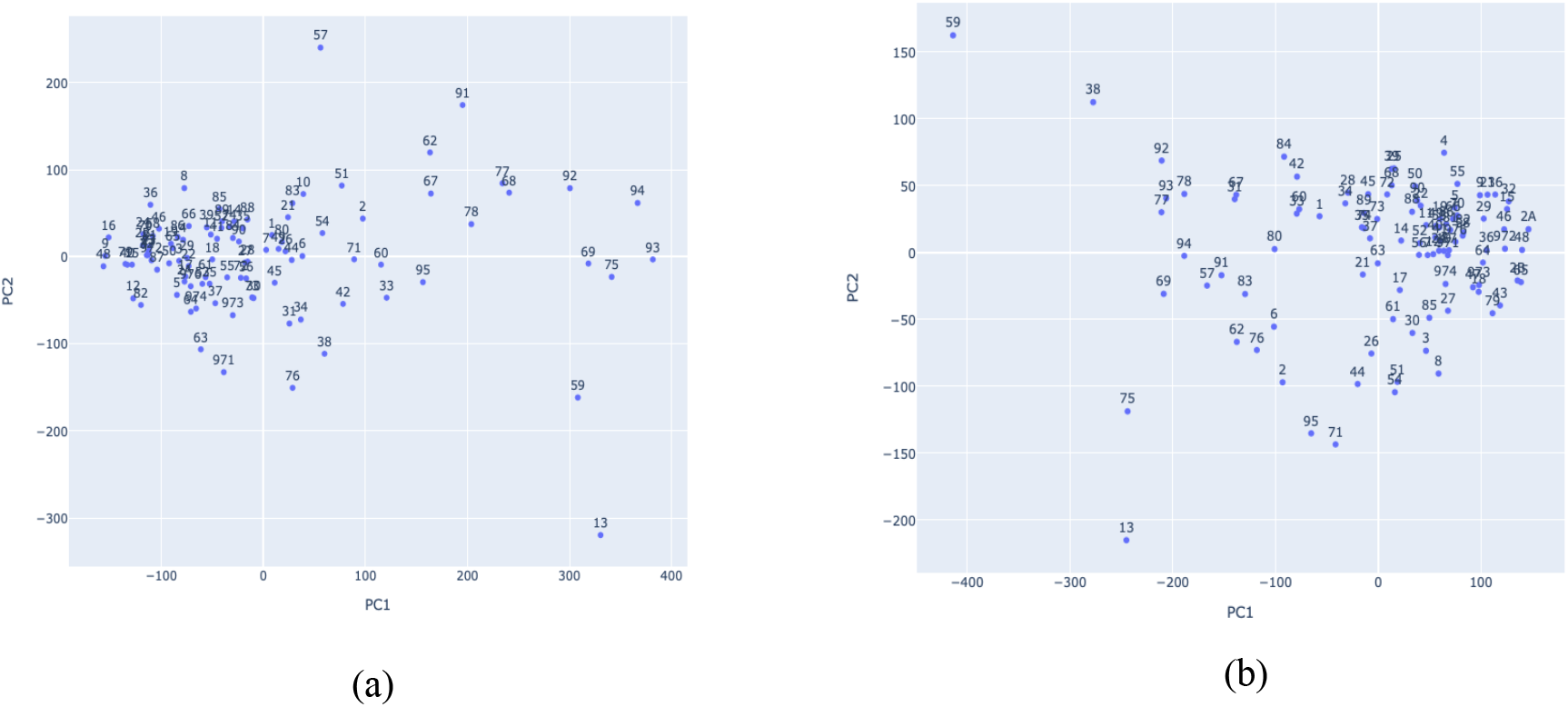
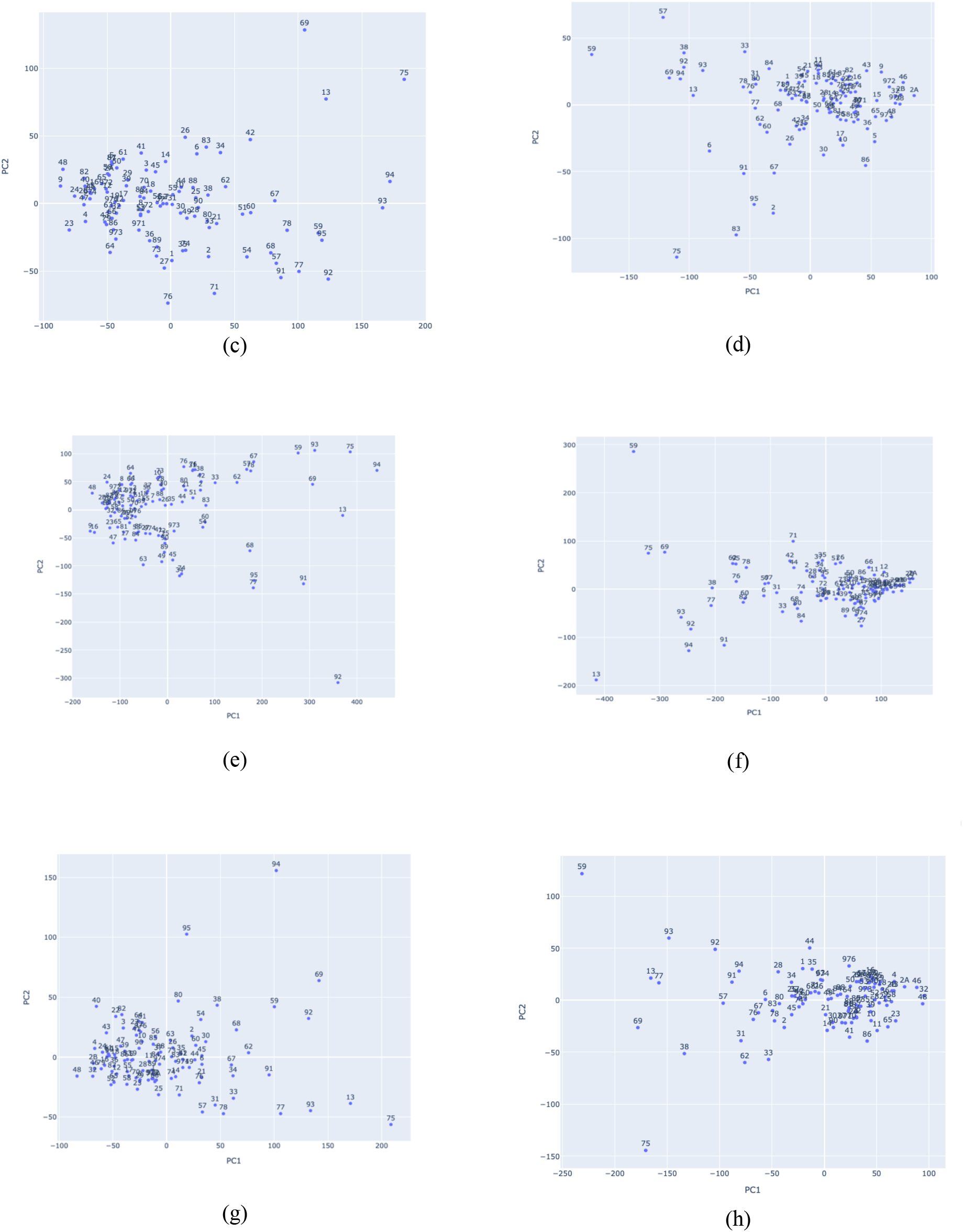
FPCA scores for different functional data before the start of vaccination (19/03/2020 to 29/10/2020) and when vaccination has started (27/12/2020 to 30/06/2021). (a) hospitalized cases, (b) hospitalized when vaccination has started, (c) ICU cases, (d) ICU cases when vaccination has started, (e) daily return home, (f) daily return home when vaccination has started, (g) daily deceased and (h) daily deceased when vaccination has started. Note that the numbering of points on the diagram are codes for each French department.

The above description shows that there is a difference between the vaccination period in France and the period when measures like lockdown, social distancing etc. were only used to control the spread of the virus despite the fact that it has been proven medically that people can be vaccinated and still be infected.

The diagrams of Figures 7a to 7h show the same shift toward positive values of PC 1.

## 4. Canonical Correlation Analysis (CCA)

Canonical correlation is an aspect of multivariate statistical analysis method that is used to simultaneously correlate several metric dependent variables and several metric independent variables measured on or observed with similar experimental units. PCA is often used for dimensionality reduction of a particular data set through linear combinations of the initial variables which maximizes the amount of variance explained by these linear combinations while CCA finds linear combinations within a data set with the goal of maximizing the correlation between these linear combinations [7].

Mathematically, it can also be expressed as two groups of n-dimensional variables X = [*x*_1_, *x*_2_, *x*_3_, … *x*_*p*_] and Y = [*y*_1_, *y*_2_, *y*_3_, … *y*_*q*_], where, 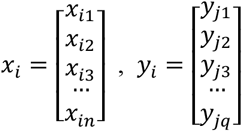.

The purpose of canonical correlation analysis is to find coefficient vectors ***a***_**1**_ = (*a*_11_, *a*_21_, …, *a*_*p*1_)^*T*^ and ***b***_**1**_ = (*b*_11_, *b*_21_, …, *b*_*q*1_)^*T*^ to maximize the correlation *β* = *corr*(*X**a***_**1**_, *Y**b***_**1**_) while *U*_1_ = *X**a***_**1**_ and *V*_1_ = *Y**b***_**1**_, linear combinations of X and Y components respectively, constitute the first pair of canonical covariates. Then, the second pair of canonical variates can be found in the same way subject to the constraint that they are uncorrelated with the first pair of variables. By repeating this procedure, *r* = *min*{*p, q*} pairs of canonical variates can be found and we will finally get two matrices A = [***a***_**1**_, ***a***_**2**_, ***a***_**3**_, … ***a***_***r***_] and B = [***b***_**1**_, ***b***_**2**_, ***b***_**3**_, … ***b***_***r***_] to transfer X and Y to canonical variates U and V following the below expression:

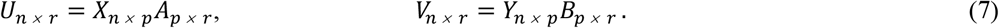

If X and Y are both centered, we can concatenate them and calculate the covariance matrix given as:

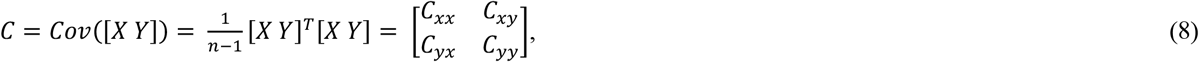

where *C*_*xx*_ and *C*_*yy*_ are within-set covariance matrices, and *C*_*xy*_= [*C*_*yx*_]^*T*^ are between-set covariance matrices. The first canonical variates ***a***_**1**_ and ***b***_**1**_ maximize the equation below:

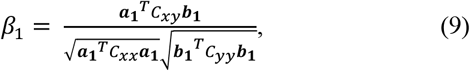

The subsequent pairs of canonical variates ***a***_***i***_ and ***b***_***i***_ (i ≥ 2) maximize:

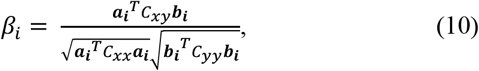

subject to the constraint:

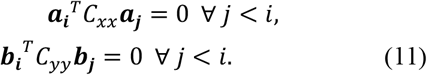

The analysis was performed in R using a package *CCA*. We present the visualization results on Figures 8 and 9 and also present the correlation scores in tabular form (see Table 3). We used the variables as presented in Table 3. X are the variables in the first column of Table 3 i.e., total number of hospitalization, daily return home, deceased and ICU cases for all departments and Y variables are the response variables described earlier as presented in the first row of Table 3.

**Figure 8.**
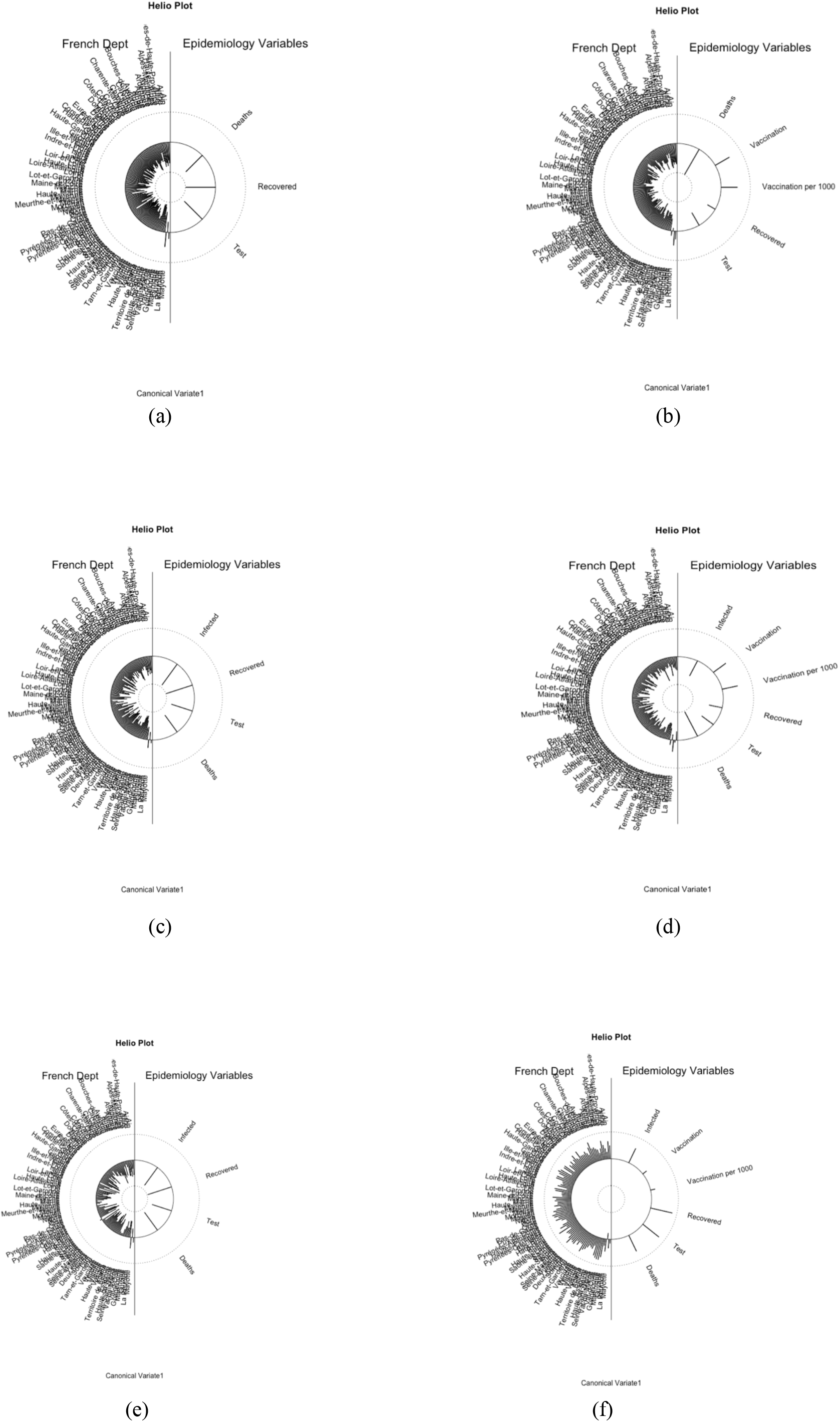
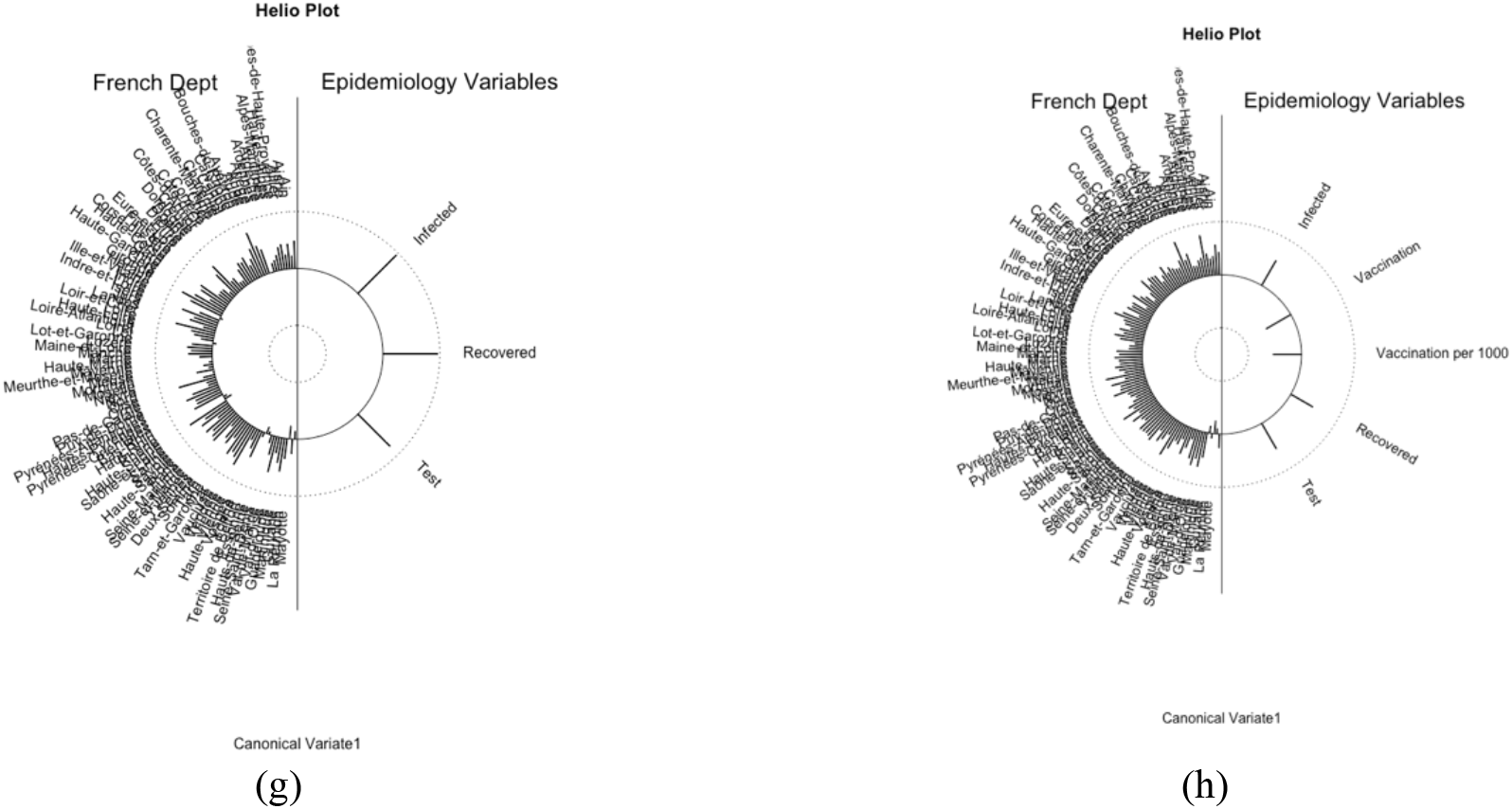
Helio plot for the correlation of French departments for (a) hospitalized cases, (b) hospitalized when vaccination has started, (c) ICU cases, (d) ICU cases when vaccination has started, (e) daily return home, (f) daily return home when vaccination has started, (g) daily deceased and (h) daily deceased when vaccination has started.

**Figure 9.**
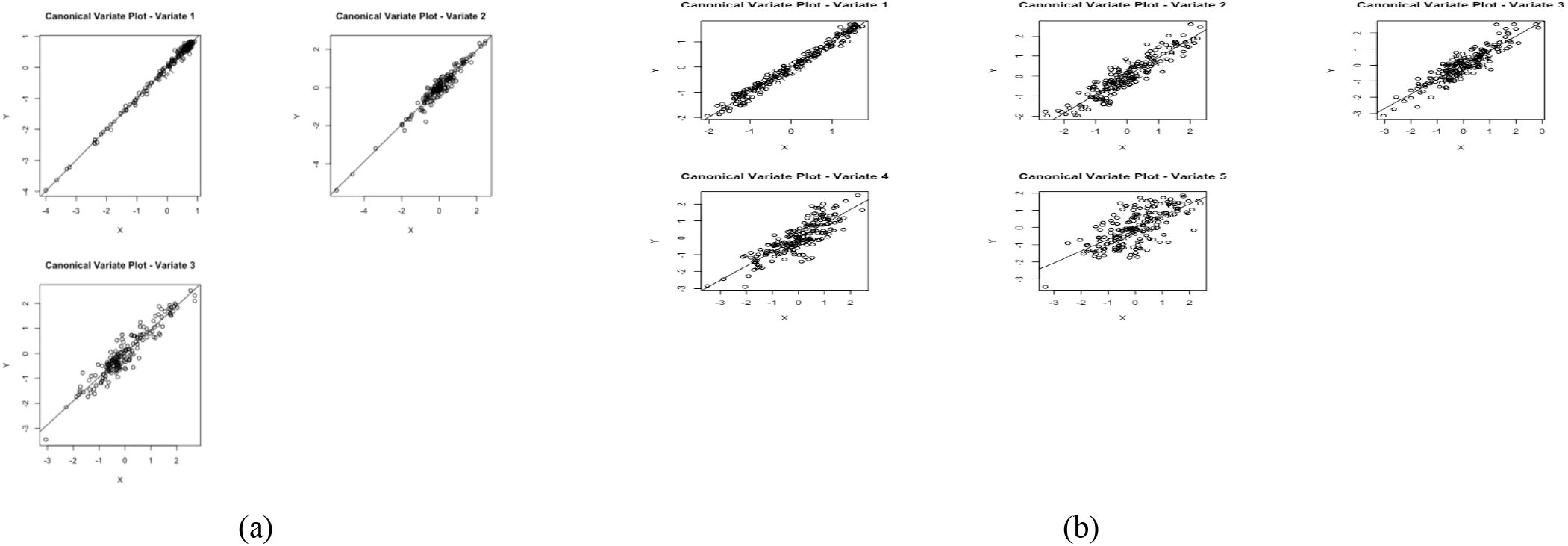
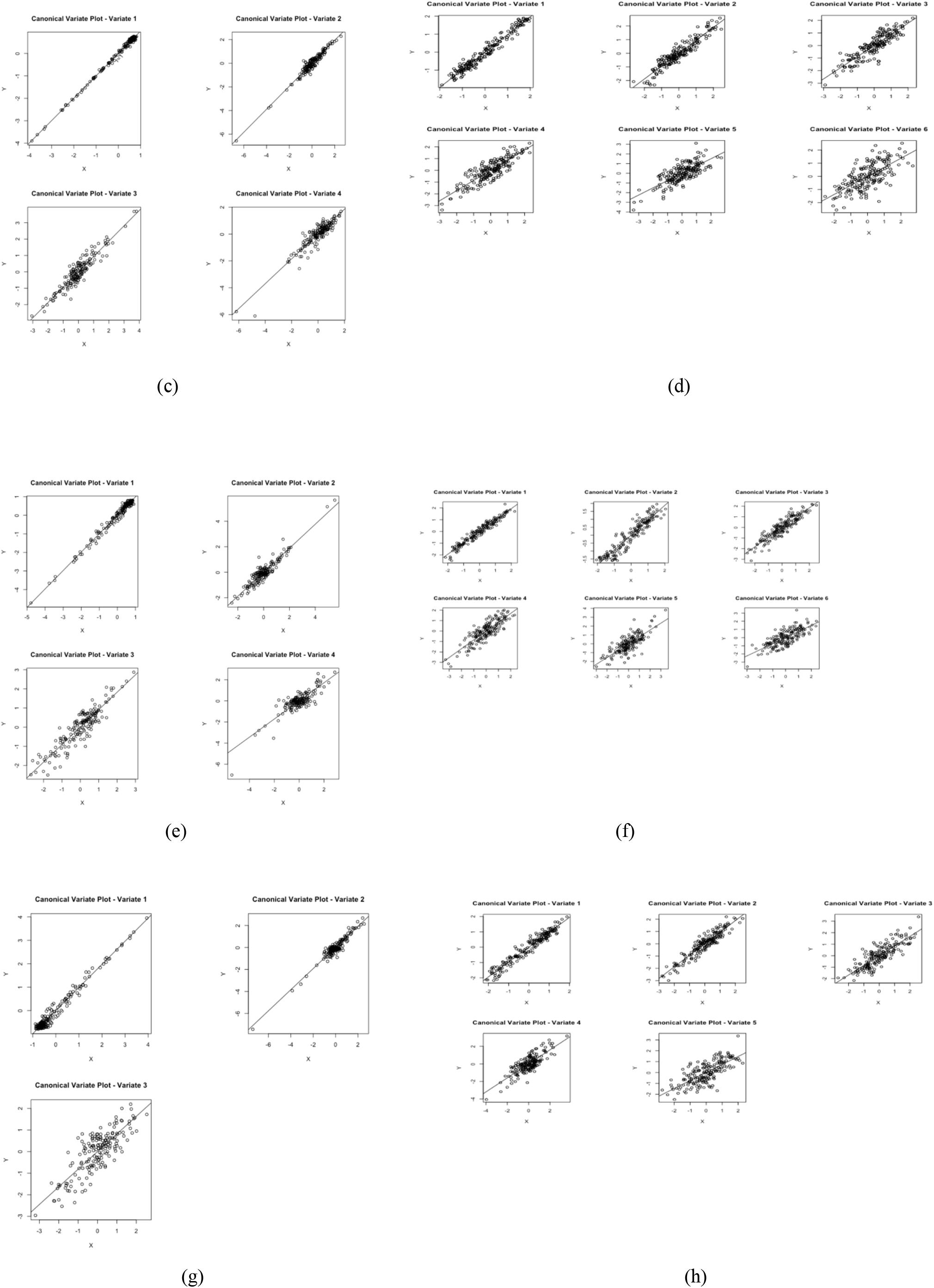
Canonical correlation visualization of (a) hospitalized cases, (b) hospitalized when vaccination has started, (c) ICU cases, (d) ICU cases when vaccination has started, (e) daily return home, (f) daily return home when vaccination has started, (g) daily deceased and (h) daily deceased when vaccination has started.

**Table 3.** Canonical correlation scores

|  | Before vaccination started |  |  |  | After vaccination has started |  |  |  |
| --- | --- | --- | --- | --- | --- | --- | --- | --- |
|  | Hospitalized | ICU | Daily return home | Daily deceased | Hospitalized | ICU | Daily return home | Daily deceased |
| Deaths | 0.996 | 0.926 | 0.859 | - | 0.989 | 0.689 | 0.745 | - |
| Recovered | 0.970 | 0.973 | 0.941 | 0.961 | 0.838 | 0.865 | 0.846 | 0.797 |
| Test | 0.950 | 0.937 | 0.911 | 0.816 | 0.685 | 0.750 | 0.776 | 0.736 |
| Vaccination | - | - | - | - | 0.924 | 0.942 | 0.939 | 0.936 |
| Infected | - | 0.998 | 0.992 | 0.987 | - | 0.980 | 0.971 | 0.970 |
| Vaccination/1000 | - | - | - | - | 0.901 | 0.885 | 0.917 | 0.841 |

The results presented in Figure 9 show the linear relations in the scatter plot as most of the variables show 95% significance level and from Table 3 there is high correlations between the variables considered. In Figure 8, the helio plot presents the relationships between the different departments in France.

Figure 9a shows hospitalized cases with p-value < 0.05 for all canonical variate, Figure 9b shows hospitalized when vaccination has started with p-value < 0.05 except the last Canonical variate with p-value= 0.88, Figure 9c shows ICU cases with p-value < 0.05 for all canonical variate, Figure 9d shows ICU cases when vaccination has started with p-value < 0.05 except the last two Canonical variate with p-value = 0.68 and p-value = 0.87 respectively, Figure 9e shows daily return home with p-value < 0.05 for all canonical variate, Figure 9f shows daily return home when vaccination has started with p-value < 0.05 except the last two Canonical variate with p-value = 0.14 and p-value = 0.34 respectively, Figure 9g shows daily deceased with p-value < 0.05 except the last Canonical variate with p-value= 0.08 and Figure 9h shows daily deceased when vaccination has started with p-value < 0.05 except the last two Canonical variate with p-value = 0.08 and p-value = 0.46 respectively.

## 5. Clustering method

The clustering of functional data is one method that statisticians are always interested in and in this Section we used the K-means and Fuzzy K-means techniques whose algorithm is already in Python *skfd.ml.clustering and FuzzyCMeans*. These methods will enable us to visualize how various departments are clustered based on our functional data and to give it the best interpretation based on their geographical location. The basic function used for the K-means clustering is a B-spline and results of our clusters are presented below. We present the result in the cluster form and also on the map of France with indication of the membership to the 3 clusters (0, 1 or 2) to get a clearer view of the result. We only presented the result for two cases (daily hospitalized and daily deceased) for the period before vaccination begins in France and two cases (daily return and ICU cases) for the period when vaccination has started in France. In Figures 11a to 11d we present the clusters (0, 1 or 2) that each French departments belongs to. The result clustered French departments outside France to the same clusters which of course are not binded with mitigation measures and rules used in departments within France. Also, departments with close proximity with Paris are in the same cluster which is the same with departments having the same trend of the pandemic prevalence as presented in Figures 11a to 11d.

**Figure 10.**
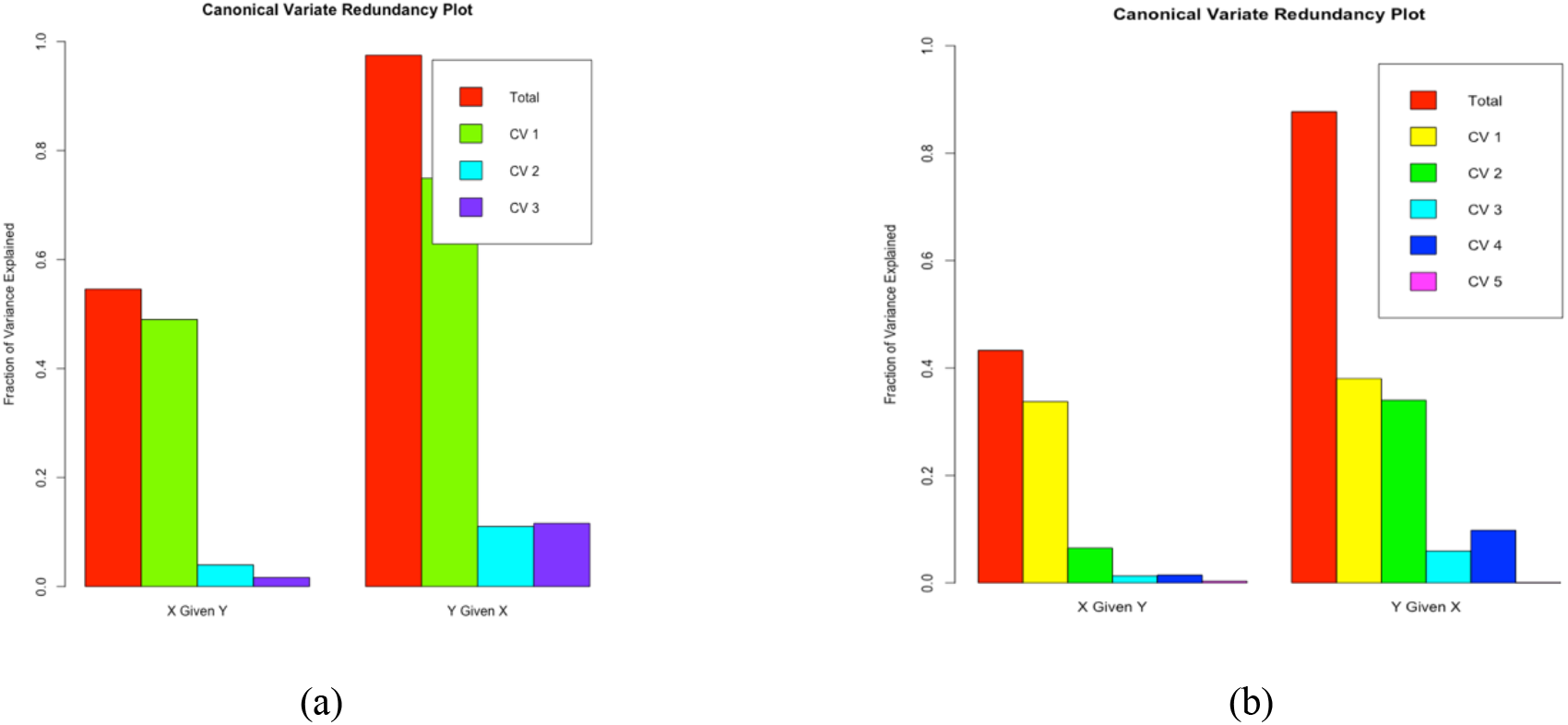
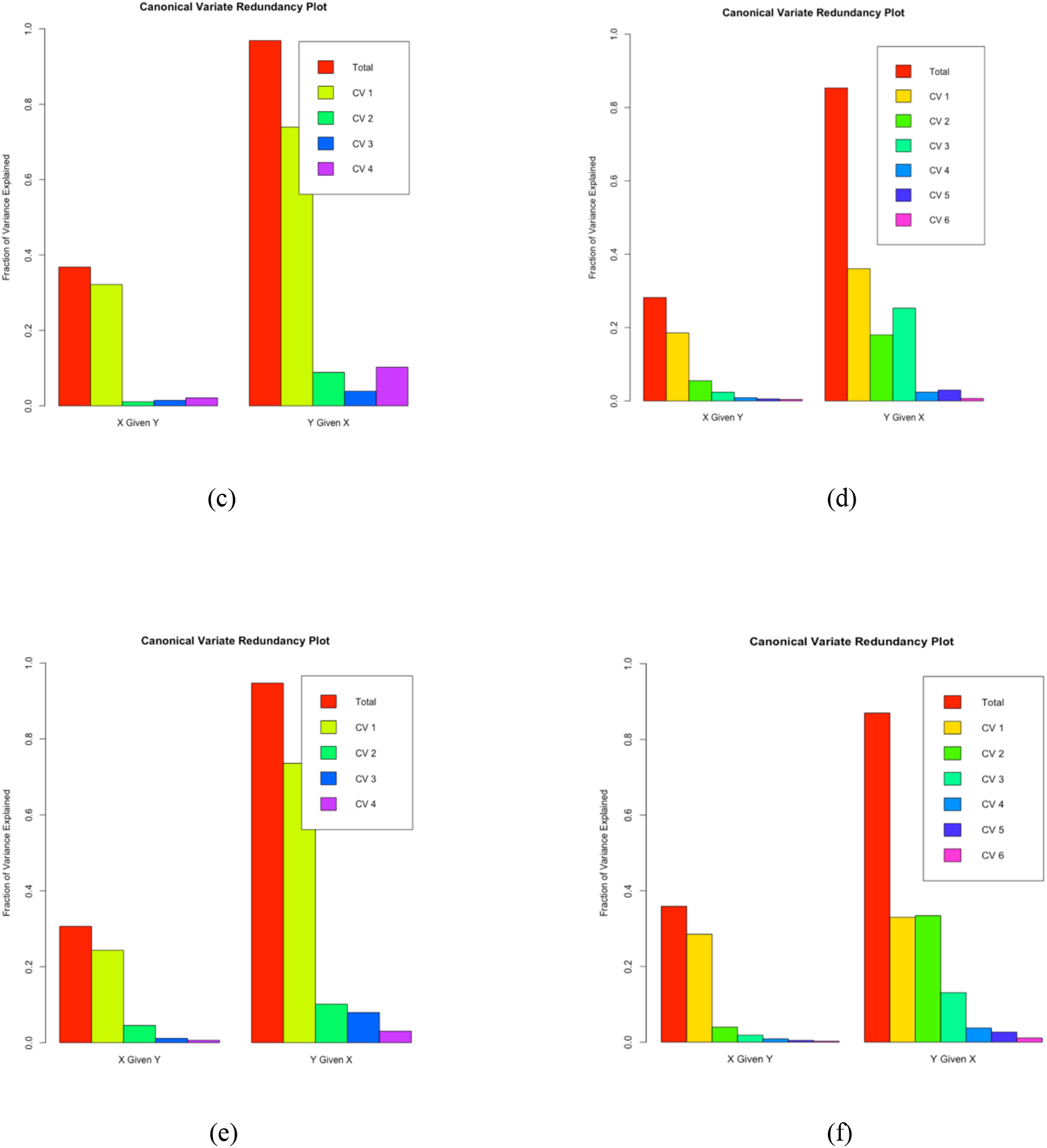
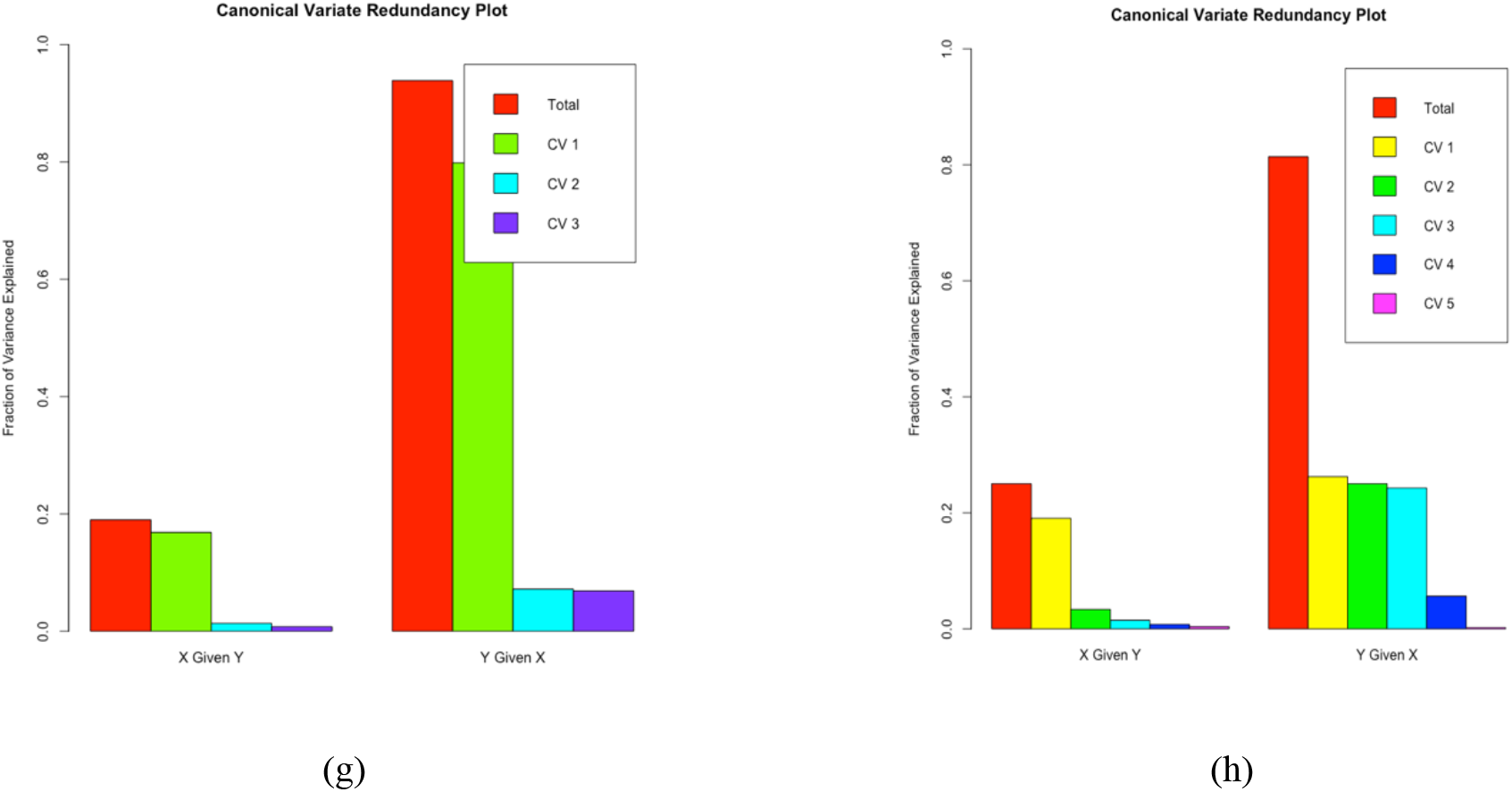
Canonical variate redundancy plot for (a) hospitalized cases, (b) hospitalized when vaccination has started, (c) ICU cases, (d) ICU cases when vaccination has started, (e) daily return home, (f) daily return home when vaccination has started, (g) daily deceased and (h) daily deceased when vaccination has started.

**Figure 11.**
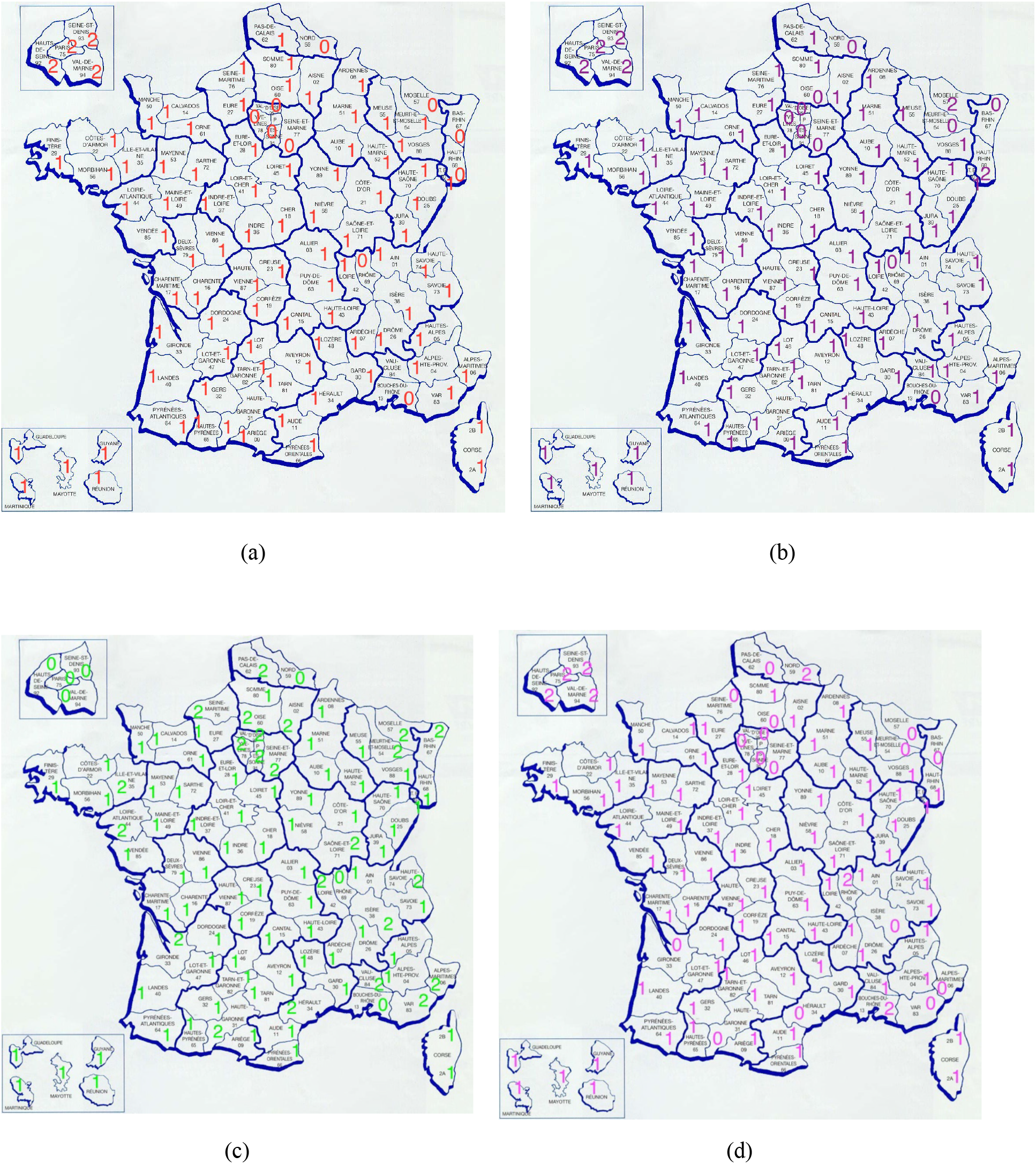
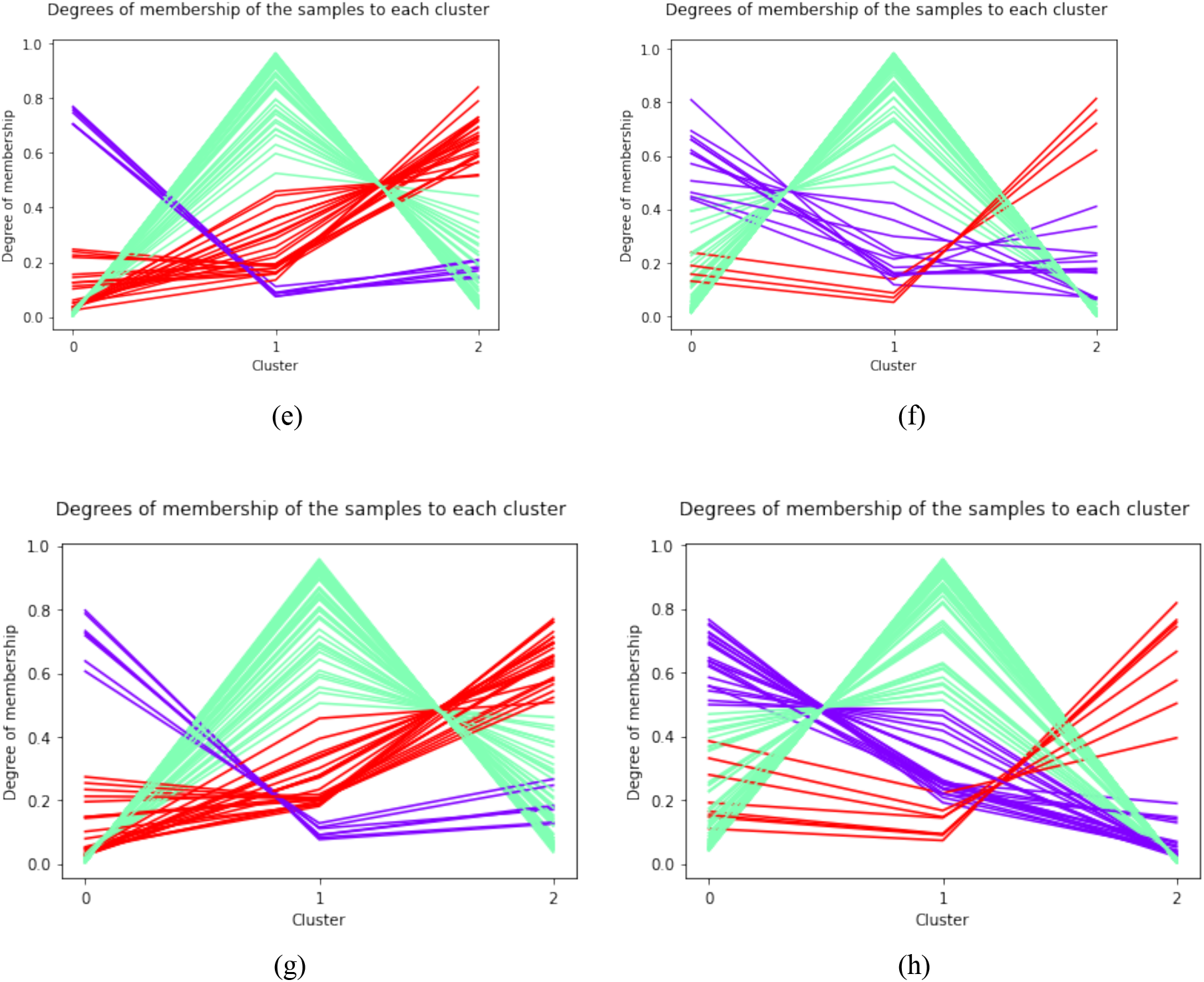
Clustering of all departments in France using K-means ((a) hospitalized, (b) daily deceased, (c) daily return home when vaccination has started and (d) ICU cases when vaccination has started) and Fuzzy K-means ((e) clusters for hospitalized, (f) clusters for daily deceased, (g) clusters for daily return home when vaccination has started and (h) clusters for ICU when vaccination has started).

## 6. Prediction

### Function to scalar linear model

In this Section we used functional linear regression model to predict two of our response variables. Let

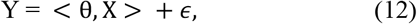

where θ is the unknown function of the model, X is a functional covariate belonging to some functional space ℍ which is endowed with an inner product <.,. >, Y is the response variable and *ϵ* is the random error term. Usually, ℍ is the space *L*^2^([*a, b*]) of square integrable functions on some real compact interval [*a, b*] and

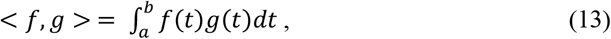

is the corresponding inner product, where the functions f, g ∈ *L*^2^([*a, b*]).

Then, we consider C= [0,1], so the equation (12) can be written as:

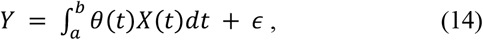

where *θ* is a square integrable function which is is defined on C and *ϵ* is a random variable such that 𝔼(*ϵ*) = 0 and 𝔼(*X*(*t*)*ϵ*) = 0. The equation 14 can be rewritten as:

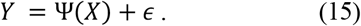

where Ψ represents the integral.

We treated the functional data (hospitalization) as a curve whose prediction is linked to a scalar (number of deaths and tests) response variable. The data considered are data before vaccination started in France and we trained 80% of the data and 20% was tested. The visualization of the results is presented in the Figure 12 and the tabular form of the numerical results can be found in Table 4. The prediction affirms the fact that the relaxation in the mitigation measures observed during the period we predicted increases the number of deaths and tests in France which is why the predicted results are a bit higher than the observed values as seen in Figure 12 and Table 4.

**Figure 12.**
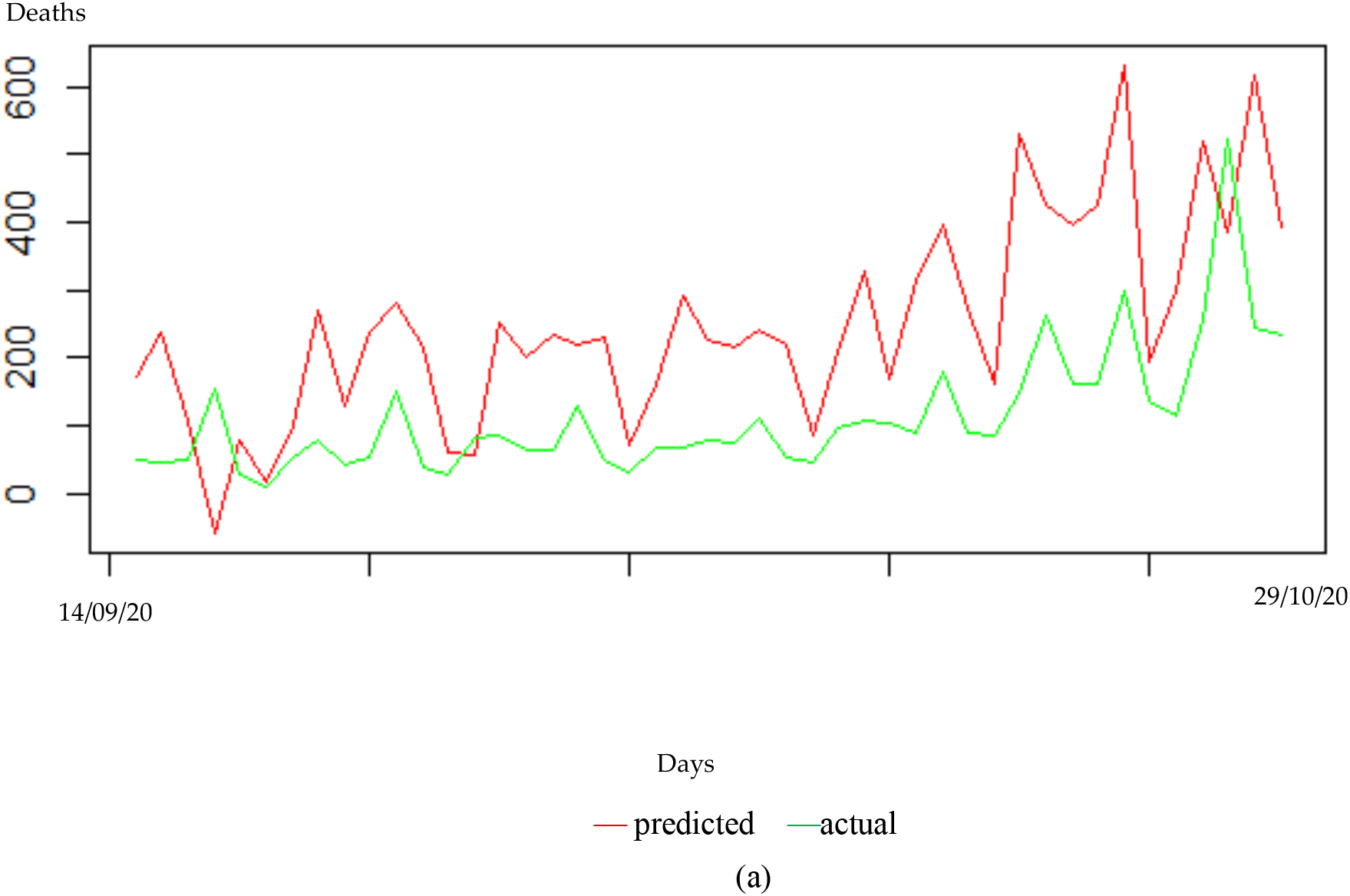
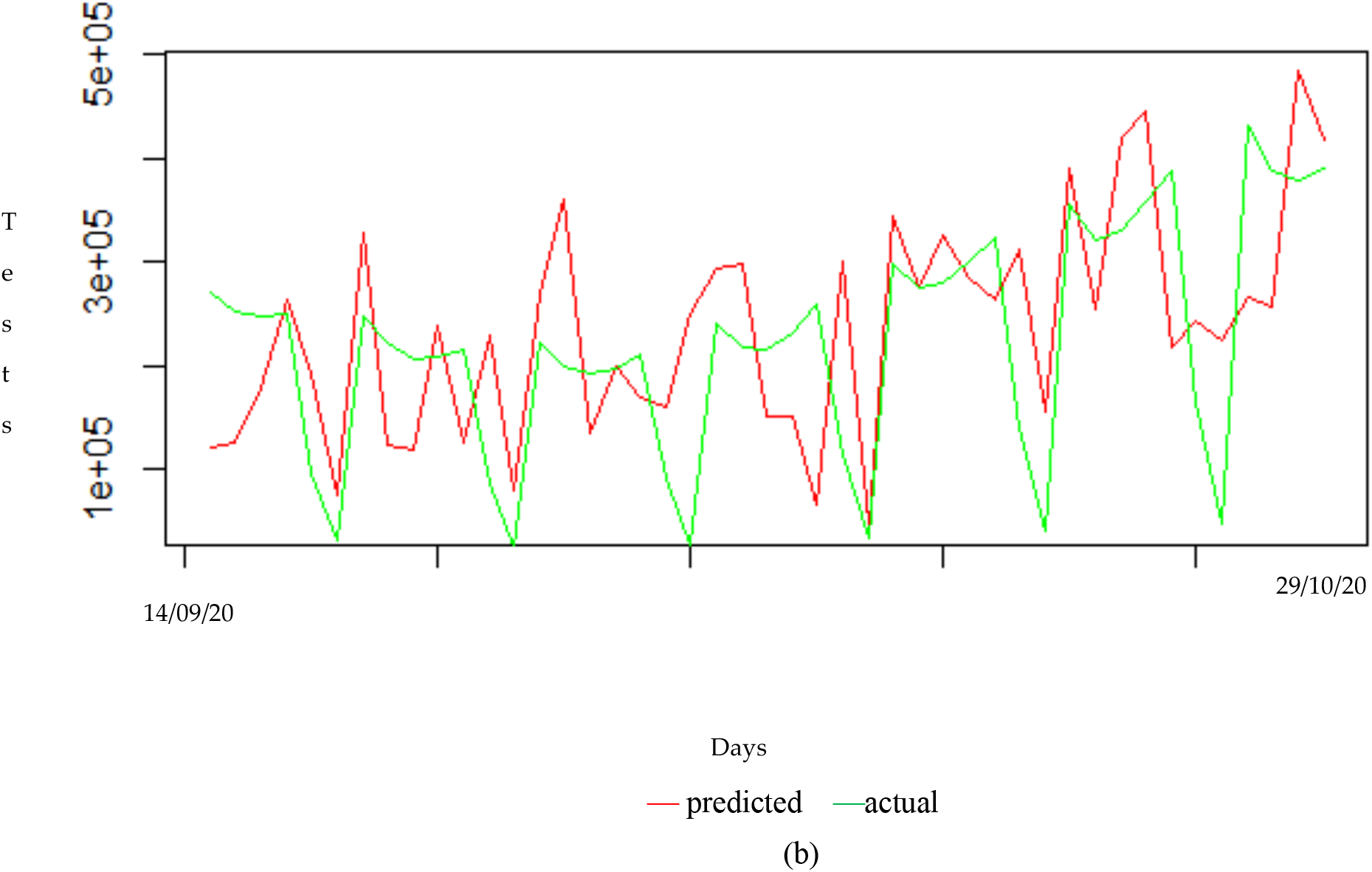
(a) Functional linear regression model prediction for number of deaths in France as response variable before vaccination begins and (b) Functional linear regression model prediction for number of tests in France as response variable before vaccination begins.

**Table 4.** Comparison of the predicted and actual values

| Deaths |  |  | Tests |  |
| --- | --- | --- | --- | --- |
| Day | Actual | Predicted | Actual | Predicted |
| 1 | 49 | 86 | 269886 | 120499 |
| 2 | 46 | 136 | 251301 | 126261 |
| 3 | 50 | 86 | 248354 | 178764 |
| 4 | 153 | 116 | 248910 | 264431 |
| 5 | 26 | 82 | 96177 | 192823 |
| 6 | 11 | 39 | 30345 | 75319 |
| 7 | 53 | 97 | 247760 | 328075 |
| 8 | 78 | 136 | 222942 | 124271 |
| 9 | 43 | 124 | 206626 | 118126 |
| 10 | 52 | 171 | 207651 | 237759 |
| 11 | 150 | 175 | 214336 | 124387 |
| 12 | 39 | 126 | 86361 | 229623 |
| 13 | 27 | 42 | 23804 | 78612 |
| 14 | 81 | 72 | 223293 | 268357 |
| 15 | 85 | 172 | 199948 | 360891 |
| 16 | 63 | 168 | 191917 | 135525 |
| 17 | 63 | 246 | 196259 | 199318 |
| 18 | 130 | 146 | 210495 | 168640 |
| 19 | 49 | 167 | 90639 | 160399 |
| 20 | 32 | 59 | 25699 | 249580 |
| 21 | 69 | 181 | 240612 | 294387 |
| 22 | 66 | 185 | 217585 | 298154 |
| 23 | 80 | 188 | 214258 | 150142 |
| 24 | 76 | 194 | 231306 | 150122 |
| 25 | 109 | 219 | 259073 | 64680 |
| 26 | 54 | 209 | 114369 | 301318 |
| 27 | 46 | 78 | 32368 | 45122 |
| 28 | 95 | 198 | 299121 | 343681 |
| 29 | 108 | 292 | 276013 | 274313 |
| 30 | 104 | 165 | 279376 | 325882 |
| 31 | 88 | 275 | 301465 | 284502 |
| 32 | 178 | 320 | 322468 | 262879 |
| 33 | 89 | 238 | 140298 | 312368 |
| 34 | 85 | 169 | 40313 | 154521 |
| 35 | 146 | 447 | 355160 | 390516 |
| 36 | 262 | 376 | 321373 | 254298 |
| 37 | 163 | 410 | 330328 | 419636 |
| 38 | 162 | 329 | 357368 | 445595 |
| 39 | 298 | 484 | 388884 | 217528 |
| 40 | 137 | 206 | 165764 | 242920 |
| 41 | 116 | 318 | 47485 | 223540 |
| 42 | 257 | 430 | 430644 | 264886 |
| 43 | 523 | 320 | 387569 | 256737 |
| 44 | 244 | 548 | 379590 | 484870 |
| 45 | 235 | 348 | 390099 | 417031 |

### Function-on-function linear model

We consider a functional input and functional output regression model where we treated y(t) as a scalar at each time t, i.e., *x*(*t*) → *y*(*t*). The functional linear model with an intercept is of the form:

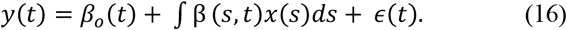

We used this method to perform a funtion-on-function linear regression on our set of functional data by using some curves to predict another set of curves while also estimating the slope *β*(*s, t*) whose results in considered cases are presented in 3D diagrams of Figure 13.

**Figure 13.**
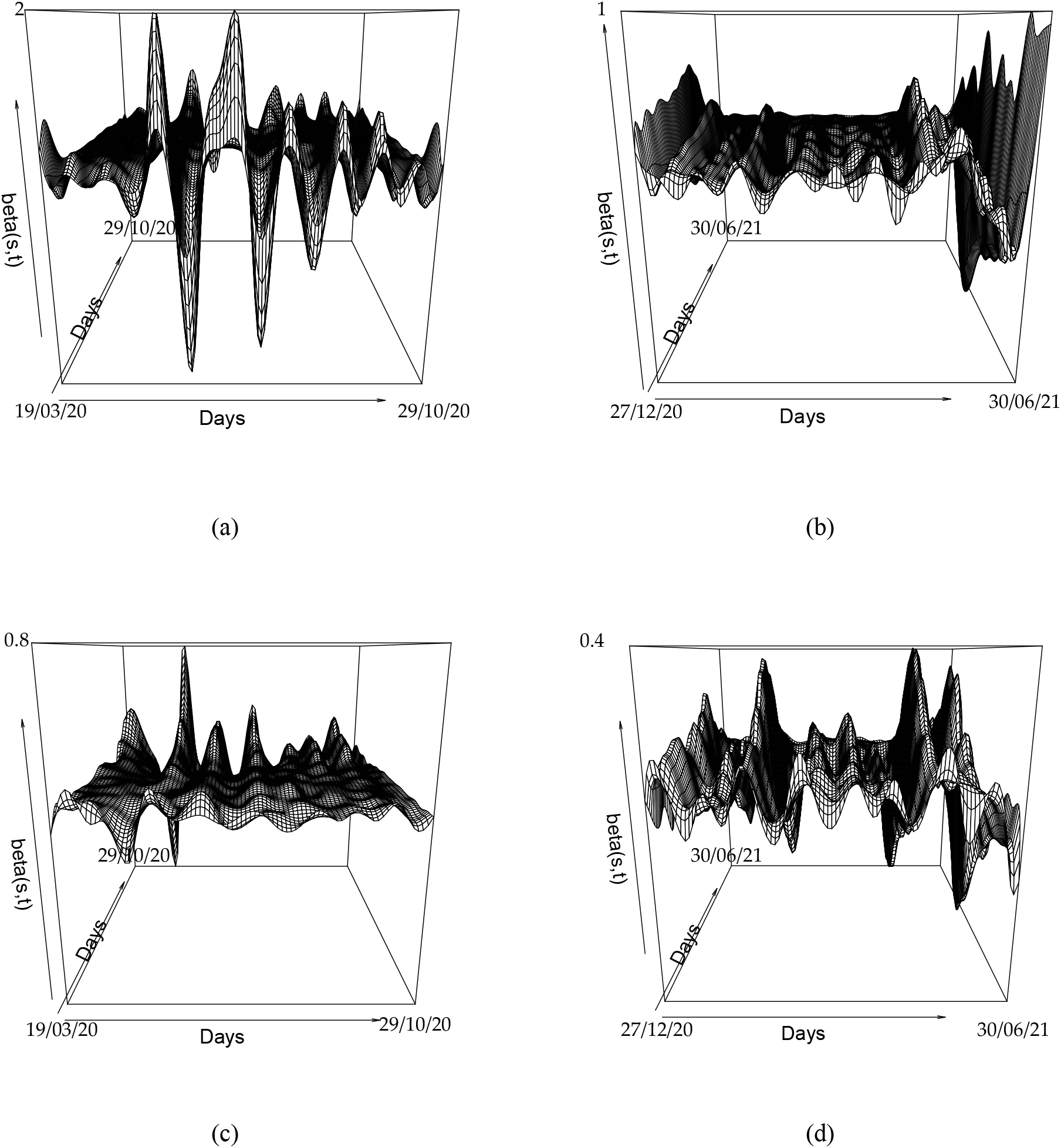
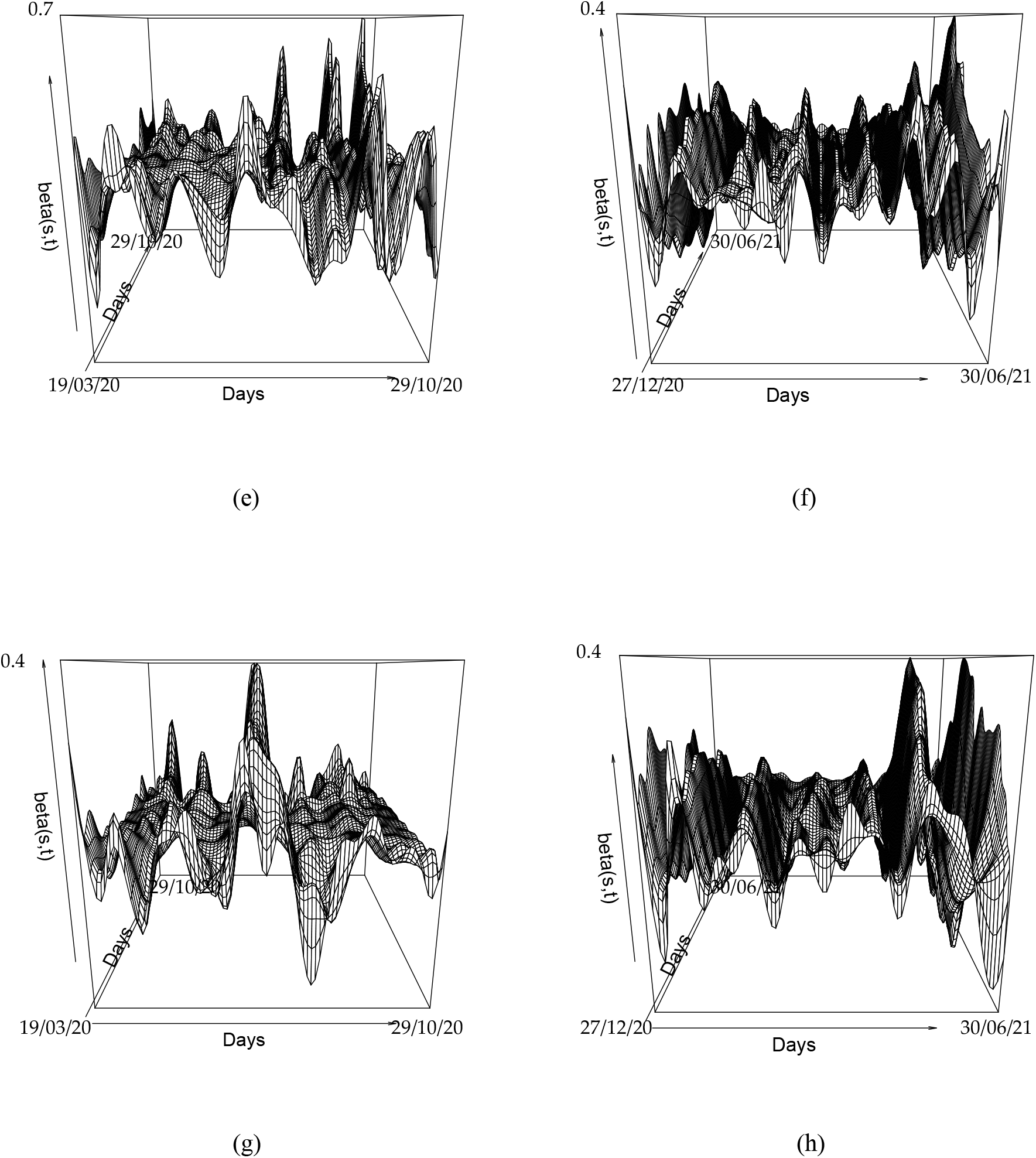
The 3D visualization of function-on-function regression for (a) hospitalized cases, (b) hospitalized when vaccination has started, (c) ICU cases, (d) ICU cases when vaccination has started, (e) daily return home, (f) daily return home when vaccination has started, (g) daily deaceased and (h) daily deaceased when vaccination has started.

Figure 13a shows hospitalized cases with the slope on the interval −2.799063 ≤ *β*(*s, t*) ≤ 1.980147, Figure 13b shows hospitalized when vaccination has started with the slope on the interval −1.501887 ≤ *β*(*s, t*) ≤ 1.076421, Figure 13c shows ICU cases with the slope on the interval −1.0733846 ≤ *β*(*s, t*) ≤ 0.8411007, Figure13d shows ICU cases when vaccination has started with the slope on the interval −0.5646148 ≤ *β*(*s, t*) ≤ 0.3661280, Figure 13e shows daily return home with the slope on the interval −0.6755000 ≤ *β*(*s, t*) ≤0.7030529, Figure 13f shows daily return home when vaccination has started with the slope on the interval −0.4333295 ≤ *β*(*s, t*) ≤0.4300995, Figure 13g shows daily deaceased with the slope on the interval −0.3277864 ≤ *β*(*s, t*) ≤0.4002531 and Figure 13h shows daily deaceased when vaccination has started with the slope on the interval −0.3284866 ≤ *β*(*s, t*) ≤0.3641679. We observed that in all these Figures in this Section, the 3D surfaces yields results whose shapes look roughly similar to the slope curve, functional predictors curve and functional response curve.

## 7. Perspectives and Conclusion

We studied in this article the best way to summarize temporal information relating to the variations of variables linked to the epidemic dynamics of COVID-19, such as hospitalized cases before and after vaccination has started, medical intensive care unit (MICU) cases before and after vaccination, daily return home cases before and after vaccination, and daily deceased before and after vaccination. Using the functional principal component analysis, it was shown that the first functional principal component well summarized the U or W shape observed for the data related to the first three principal components. This discovery confirms the importance of this first component for the explanation and the qualitative prediction from the observed data. The influence of vaccination is visible, because the U or W shape is attenuated after vaccination, and does not come close to the shapes observed for seasonal influenza [16]. The subsequent functional principal components have poor predictive power, but the second component clearly shows the reducing influence of vaccination on all epidemic variables. A further, more in-depth study could undoubtedly show the predictive nature of this second component on the future success of a vaccination policy, by comparing different countries with different vaccination rates and by quantifying the phase of descent of the curves of the second component (for example by its slope at the second inflection and by the value of its minimum).

## Data Availability

All data is available in public databases

## APPENDIX A

**Figure 14.**
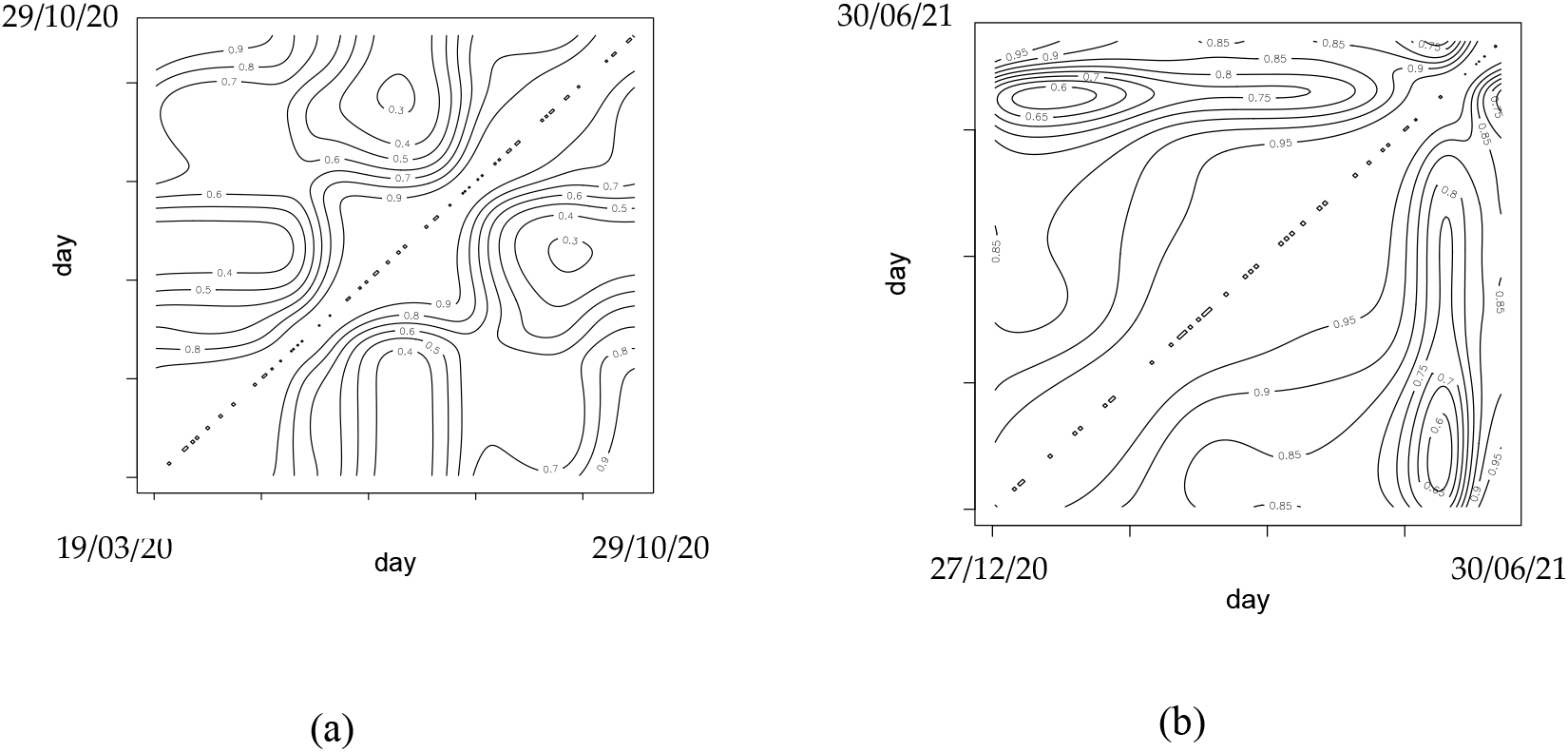
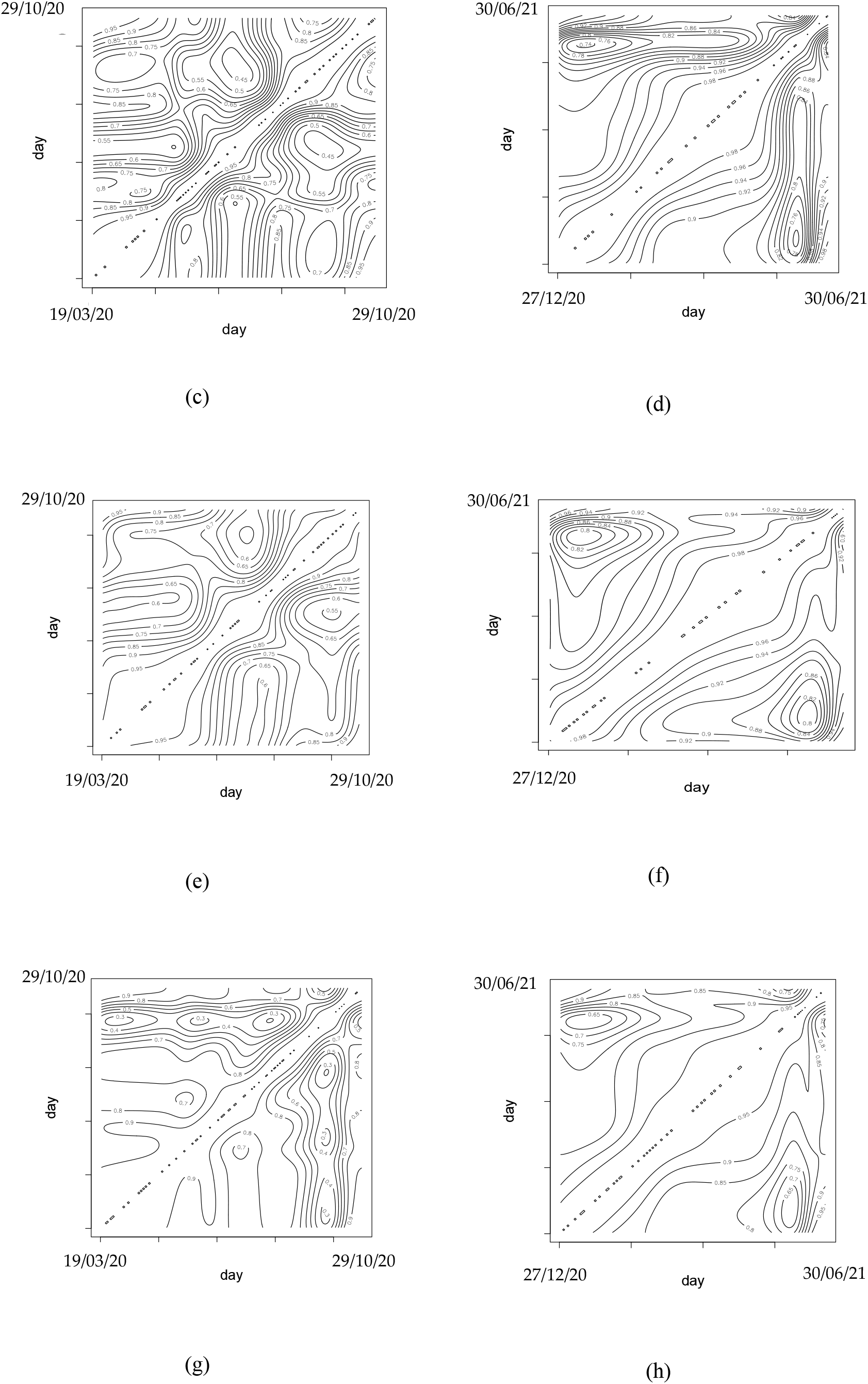
Correlation coefficients between all French departments contour plot. (a) hospitalized cases, (b) hospitalized when vaccination has started, (c) ICU cases, (d) ICU cases when vaccination has started, (e) daily return home, (f) daily return home when vaccination has started, (g) daily deceased and (h) daily deceased when vaccination has started.

## APPENDIX B

**Figure 15.**
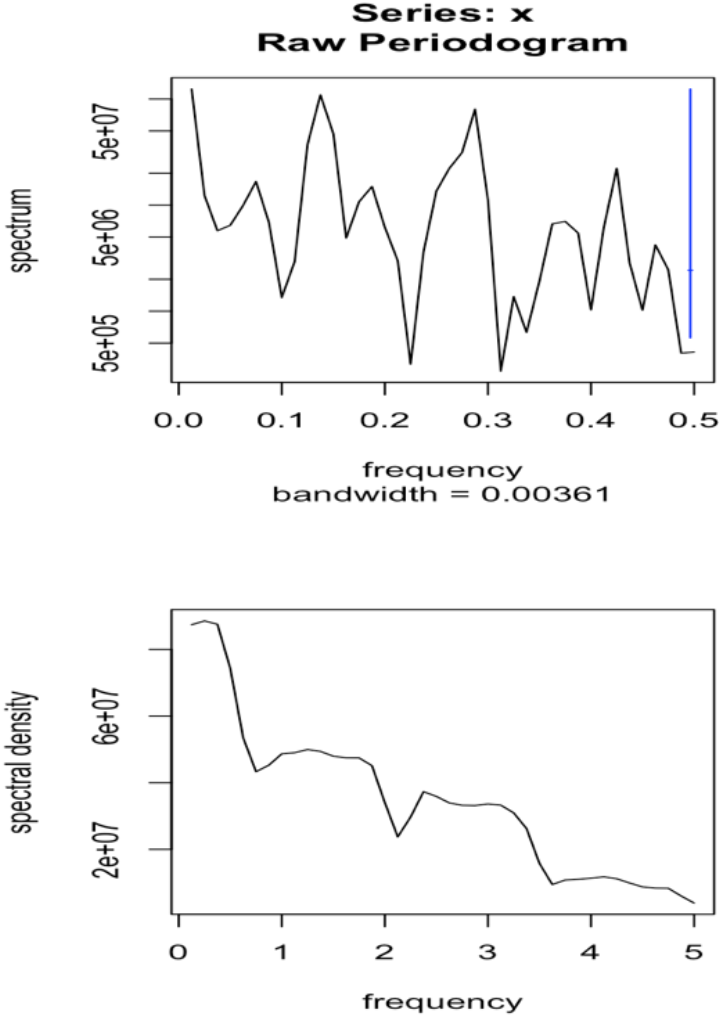
Spectral analysis of daily new cases between 01/05/2021 to 15/07/2021 in France.

## Author’s contribution

Conceptualization, J.D.; K.O. and M.R.; methodology, J.D.; K.O.; F.I. and M.R.; software, K.O. and F.I.; validation, J.D.; K.O. and M.R.; formal analysis, K.O. and F.I.; investigation, J.D. and M.R.; resources, J.D.; data curation, K.O. and F.I.; writing—original draft preparation, K.O.; writing—review and editing, J.D. and K.O; visualization, K.O. and F.I.; supervision, J.D. and M.R; project administration, J.D. and M.R. All authors have read and agreed to the final version of the manuscript.

## Conflict of interest

Authors declare no conflict of interest.

## Funding

No specific funding was received for this research.

## Acknowledgments

The authors wish to acknowledge the Petroleum Technology Development Fund (PTDF) Nigeria doctoral fellowship in collaboration with Campus France Africa Unit.

## Notes

### Competing Interest Statement

The authors have declared no competing interest.

### Funding Statement

No external funding was received

### Author Declarations

All data comes from public databases

